# Neuroactive metabolites and bile acids are altered in extremely premature infants with brain injury

**DOI:** 10.1101/2023.05.17.23290088

**Authors:** Manuel Pristner, Daniel Wasinger, David Seki, Katrin Klebermaß-Schrehof, Angelika Berger, David Berry, Lukas Wisgrill, Benedikt Warth

## Abstract

The gut microbiome has been associated with pathological neurophysiological evolvement in extremely premature infants suffering from brain injury. The exact underlying mechanism and its associated metabolic signatures in infants are not fully understood. To decipher metabolite profiles linked to neonatal brain injury, we investigated the longitudinal fecal and plasma metabolome of 51 extremely premature infants using LC-HRMS-based untargeted metabolomics. This was expanded by an investigation of bile acids and amidated bile acid conjugates in feces and plasma by LC-MS/MS-based targeted metabolomics. The resulting data was integrated with 16S rRNA gene amplicon gut microbiome profiles as well as patient cytokine, growth factor and T-cell profiles. We identified an early onset of differentiation in neuroactive metabolites and bile acids between infants with and without brain injury. We detected several bacterially-derived bile acid amino acid conjugates and secondary bile acids in the plasma already three days after delivery, indicating the early establishment of a metabolically active gut microbiome. These results give new insights into the early life metabolome of extremely premature infants.

## 1 Introduction

Extremely premature infants are born before 28 weeks of gestation and represent a highly vulnerable patient cohort. While the overall survival rate of premature infants has improved in the last decades, a significant number of infants still suffer from severe morbidities and life-long neurodevelopmental impairment.^1^ Extremely premature infants are born during a critical window of human fetal development, leaving them prone to early physical, microbial, and environmental exposures. Within this third trimester window, neurophysiological maturation and development appears to be highly influenced by postnatal factors, especially by the developing gut microbiome. We recently described gut-immune-brain interactions in a clinical cohort of 60 extremely premature infants, and found that *Klebsiella* overgrowth is associated with pro-inflammatory immunological imprinting and neonatal brain injury.^2^ Therefore, early gut-immune-mediated and host-microbe signaling might have implications for early development ^3^.

The gut-immune-brain axis represents a complex bidirectional communication system comprised of various neural pathways with neuropeptides, cytokines and hormones as signaling molecules. The most direct route that connects the gut and the brain is the cranial vagus nerve. The gut is innervated throughout by its celiac and hepatic branches. Another important communication pathway between gut microbiota and the CNS is the enteric nervous system.^4^ Recent advances in understanding of the gut-immune-brain axis showed that microbes play a significant role in signal transduction, which, in return, can modulate the behavior of the host.^5, 6^ With host-affecting bacterial metabolites such as bile acid metabolites, short chain fatty acids (SCFAs), amino acid metabolism, serotonin, catecholamines, and gamma-aminobutyric acid (GABA)^7^, gut bacteria can be associated with functions and diseases of the central nervous system (CNS).^8, 9^ The synthesis of phenylalanine (Phe), tryptophan (Trp) and tyrosine (Tyr) via the shikimate pathway and subsequent catabolism results in formation of neurotransmitters l-3,4-dihydroxyphenylalanine (L-Dopa), dopamine, epinephrine and norepinephrine.^8^ Gut bacteria, on the other hand, can transform the said amino acids into phenolic compounds such as *p*-cresol. This metabolite is further metabolized in the liver to *p*-cresol sulfate, which is implicated in various neurodevelopmental disorders.^8^ The Trp-derived indole and its derivatives indole acetic acid, indole propionic acid and tryptamine, as well as kynenine and quinolinate generated through TRYP6 induce neuroactivity and potentially limit the TRP availability for the neurotransmitter (e.g. serotonin) synthesis.^10^ Other metabolites implicated in this respect are indoxyl sulfate and indole-3-lactic acid.^11^ The latter was also associated with anti-inflammatory effects in necrotizing enterocolitis (NEC) in premature infants.^12^

Recently, it has been shown that the communication between the bile system and gut microbiota is bidirectional in nature, potentially involving neural modulation. The primary role of bile acids is to facilitate the absorption of dietary fat and fat-soluble vitamins from the gut lumen.^13^ Bile acids are synthesized from cholesterol and conjugated with taurine or glycine. In humans, *de novo* synthesized primary bile acids are cholic acid (CA) and chenodeoxycholic acid (CDCA). Upon exercising their role in lipid digestion, most bile acids are recycled back to the liver *via* the ileum in active uptake through apical sodium bile acid transporters or through passive diffusion in the jejunum and colon as a part of the enterohepatic cycle. Various members of the gut microbiota (e.g., *Lactobacillus, Bifidobacterium, Enterococcus, Clostridium,* and *Bacteroides*) express bile salt hydrolase enzymes that deconjugate bile acids from taurine and glycine, which would render them less susceptible to efficient reabsorption in the enterohepatic cycle.^14, 15^ Deconjugation and dehydroxylation, oxidation and epimerization of bile acids by gut microbiota are the main mechanisms for the formation of secondary bile acids, such as deoxycholic acid (DCA), lithocholic acid (LCA), hyodeoxycholic acid (HDCA) and ursodeoxycholic acid (UDCA).^16^ They can be absorbed into the colon by passive diffusion and transported to the liver, where a repeated conjugation with glycine and taurine can occur, followed by secretion into the intestinal tract.^17^ A particularly important role of bile acids is their relation to inflammatory responses. While hydrophobic bile acids may act as pro-inflammatory signals, hydrophilic bile acids were shown to have the opposite effect. Moreover, deficits in luminal levels of bile acids were correlated to gut bacterial overgrowth, intestinal inflammation, and tissue damage.^18, 19^ Recent evidence suggested that bile acids might directly affect the brain.^20^ Bile acids act as steroid hormones, interacting with receptors like the farnesoid X receptor (FXR), sphingosine-1-phosphate receptor 2 (S1PR2) and Takeda G protein-coupled receptor 5 (TGR5).^21^ Indirectly, the influence over the central nervous system could be exerted *via* the gut-immune-brain axis, following the FXR/FGF19 cascade, activated primarily by cholic acid and chenodeoxycholic acid.

The investigation of this broad variety of different chemical compounds affecting the brain-gut axis, poses a challenge and requires a comprehensive analytical approach. High resolution mass spectrometry (HRMS) based untargeted metabolomics is an emerging and powerful discipline that enables the global analysis of endogenous and exogenous metabolites, thereby revealing information about progression, causes and potential biomarkers of diseases.^22, 23^ Metabolomics investigations of extremely premature infants have been limited to only a few publications, investigating only a single sample matrix or using a less comprehensive approach.^24–27^

We investigated the fecal and plasma metabolome in longitudinally-collected samples obtained from the PreMiBraIn cohort^2^ of extremely premature infants by untargeted LC-HRMS and targeted LC-MS/MS based metabolomics. The resulting data was then integrated with 16S rRNA gene amplicon gut microbiome profiles as well as patient cytokine, growth factor and T-cell profiles. Our goal was to decipher links between endogenous metabolites, the gut microbiome and brain injury in extremely premature infants. This work represents the first and most comprehensive untargeted metabolomics investigation of combined fecal and plasma samples from extremely premature infants with brain injury thus far. We found an early onset of differentiation in neuroactive metabolites and bile acids in extremely premature infants with subsequent brain injury diagnosis. In addition, our newly established targeted-approach detected numerous bacterial-derived amidated bile acid conjugates. Furthermore, we found secondary bile acids already three days after delivery in the plasma, indicating the establishment of a metabolically-active gut microbiome in the early postnatal phase.

## 2 Results

### 2.1 Time-dependent development is the dominating factor in overall differences in plasma and fecal metabolome

From a cohort of 51 extremely premature infants (gestational age <28 weeks, birth weight < 1000 gram), 120 plasma samples at five time points (day 3, day 7, day 28, correction week 32, term equivalent age) and 75 fecal samples at three time points (day 7, day 28, term equivalent age) were collected. The cohort consisted of two patient groups, one with age adequate findings or mild brain injuries based on MRI findings at term equivalent age, which were termed the control group (CTR, n=36), and one group with pathological and severe brain injuries, termed the pathological group (PAT, n=15).

A visual summary of the patient cohort, sample composition and experiments is illustrated in Fig. 1A. Detailed information about the patient cohort, sampling procedures, details of previous T-cell, cytokine, growth factor, 16S rRNA gene amplicon analysis and determination of brain damage is described in detail in our previous publication.^2^ For characterization of the gut and plasma metabolome, each sample was analyzed by LC-HRMS in an untargeted manner using two complementary separation methods (RP & HILIC) in positive and negative polarity mode, to ensure the coverage of a broad spectrum of compounds and chemical classes. Reversed phase chromatography (RP) was used for the separation of non-polar compounds including fatty acids, steroids, peptides and bile acids. HILIC chromatography is suitable for the measurements of moderately polar to polar compounds like amino acids, nucleotides and sugars.

**Fig. 1.**
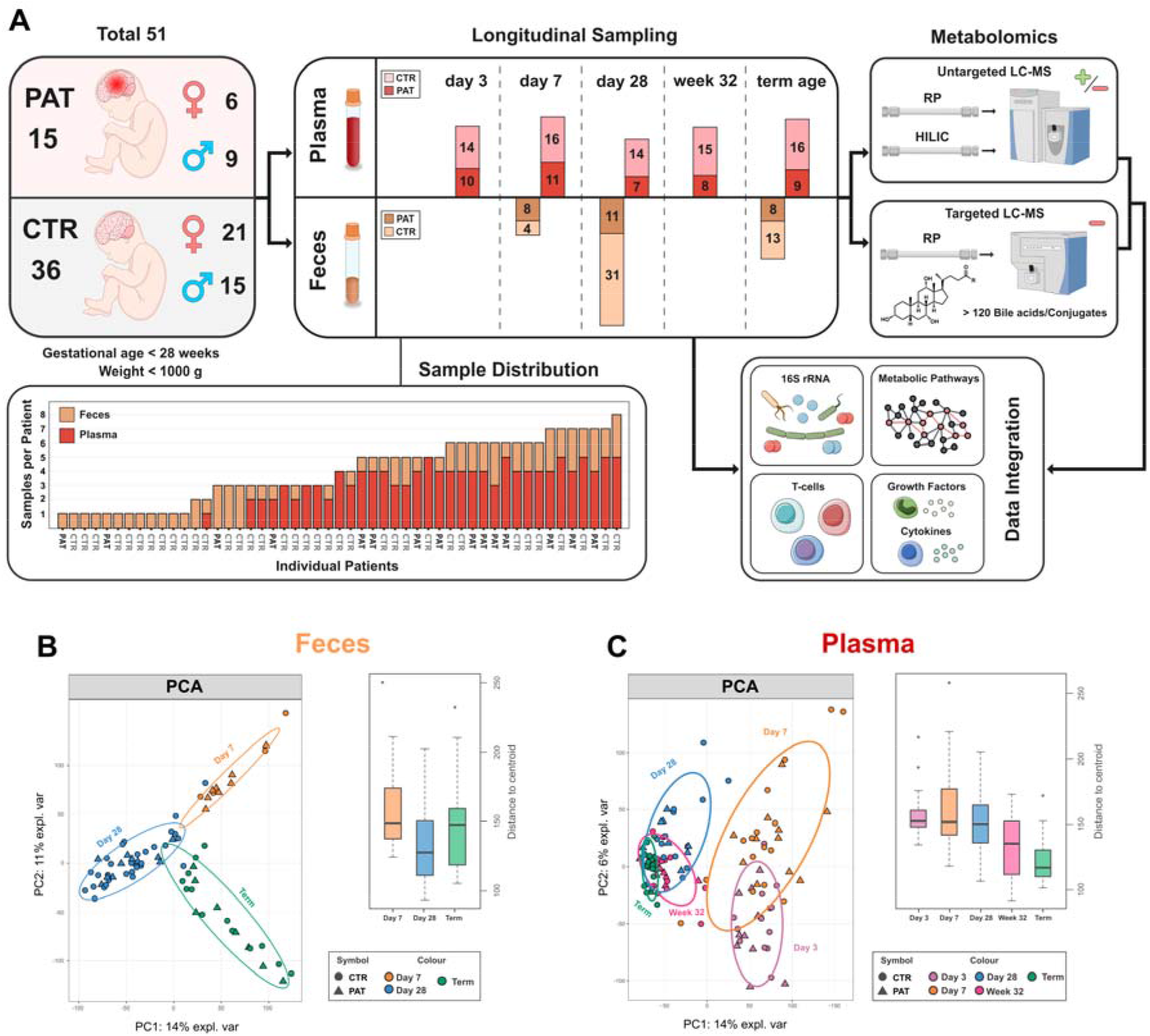
(A) Visual summary of the patient cohort, sample composition and experiments done. (B) PCA plots and boxplots of the distance to the centroid of the combined (RP + HILIC/negative + positive polarity) data obtained from the untargeted LC-HRMS experiments of all sampling time points of fecal samples. (C) PCA plots and boxplots of the distance to the centroid of the combined (RP + HILIC/negative + positive polarity) data obtained from the untargeted LC-HRMS experiments of all sampling time points of plasma samples.

For a general overview of the differences in the plasma and fecal metabolome between the groups over time, a principal component analysis (PCA) was performed on the combined data of the untargeted LC-HRMS experiments. Generally, the metabolome of the pathological and the control group exhibited no group-specific clustering in the PCA, neither in the plasma nor in the feces. But there was clearly clustering between sampling time points, as seen in plasma and feces (Fig. 1B & 1C). This finding was supported by a permutational multivariate analysis of variance (PERMANOVA) which found that time point was a significant factor (plasma: p < 0.001, feces: p < 0.001; variation explained by experimental group: plasma = 21 %; feces = 19%) while group affiliation was not deemed to be significant. This indicates that metabolic changes specific for the differentiation between the pathological group and control group (PERMANOVA variation explained by experimental group: plasma < 1 %, feces < 1 %) might be only based on a subset of metabolites, and in general individual factors dominate the differences in the metabolome between patients (PERMANOVA variation explained by patient ID: plasma = 30%, feces = 55 %). In addition, the dispersion of samples at the different sampling time points in plasma (average euclidean distance to centroid: day 3 = 157.0, day 7 = 164.5, day 28 = 151.8, week 32 = 133.6, term = 122.6) (Fig. 1C) and feces (average euclidean distance to centroid: day 7 = 160.1, day 28 = 132.9, term = 146.1) (Fig. 1B) was analyzed. The difference in dispersion based on time point was deemed significant by analysis of variance (ANOVA) in both plasma and feces (plasma: p < 0.001, feces: p = 0.017). The plasma metabolome in the early time points, especially on day 7, displayed a less tight clustering and seemed to exhibit more spread compared to term equivalent age (Fig. 1C), indicating a higher susceptibility to variation during earlier periods of infant development. At term equivalent age, a tight clustering between samples was seen, indicating a general stabilization and convergence of the plasma metabolome over time. The opposite trend was observed in the fecal samples (Fig. 1B), where a slightly tighter clustering was shown at day 28 accompanied by a trend to spread out at term equivalent age, indicating a diversification of the gut metabolome over time and possibly also the gut microbiome.^28^

In order to put the data from the untargeted LC-HRMS experiments into a biological context, we used a combination of mummichog^29^, a tool for pathway enrichment based on putative compound annotation, and a gene set enrichment analysis (GSEA) based approach. The web application MetaboAnalyst (5.0)^30^ and its “MS Peaks to Pathways” tool, based on putative metabolite annotation, was employed for this task. Already at the earliest sampling time points, day 3 for plasma and day 7 for feces, significant changes in several metabolic pathways between the pathological and control group were observed (Fig. 2A & 2B).

**Fig. 2.**
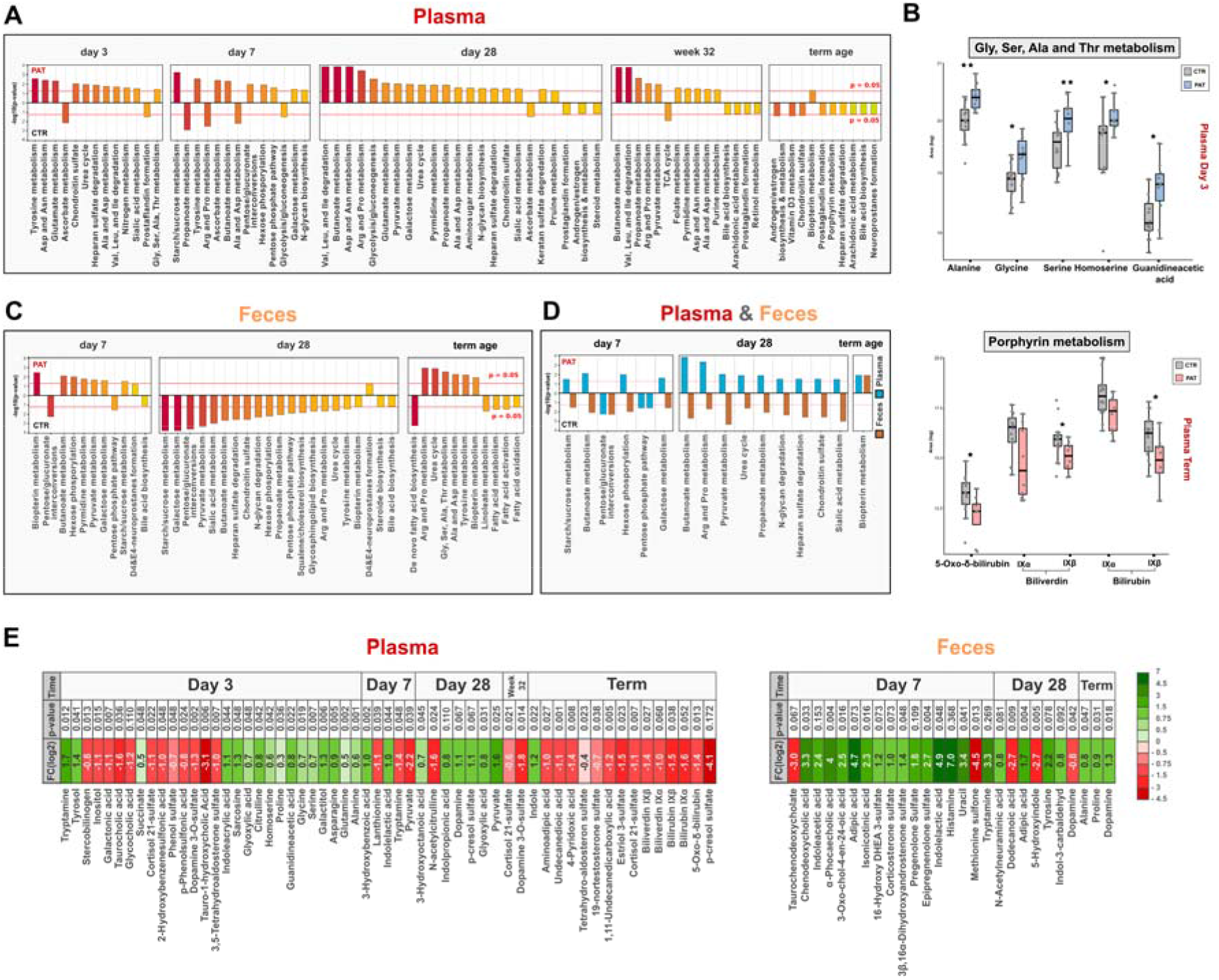
(A) Barplot plots showing the enriched metabolic pathways for plasma and their respective combined p-values obtained from the “MS Peaks to Pathways” tool. (B) Box plots of identified metabolites of amino acid and porphyrin metabolism in plasma at day 3 and term equivalent age. (C) Barplots showing the enriched metabolic pathways for feces and their respective combined p-values obtained from the “MS Peaks to Pathways” tool. (D) Enriched metabolic pathways intersection of plasma and feces at their respective time points. (E) Overview of selected metabolites and their change between experimental groups (PAT/CTR, positive FC (log2) indicates an increase in abundance in the pathological group) at different sampling time points (*additional details in SI Tab. S2)*.

In feces, at all three time points a significant or near-significant enrichment was found in biopterin metabolism (day 7: p = 0.004, day 28: p = 0.055, term: p = 0.010) (Fig. 2B). Being a cofactor for aromatic amino acid hydroxylases, biopterin is involved in the synthesis of neurotransmitters including dopamine and serotonin.^31^ Various pathways involved in carbohydrate metabolism were significantly modulated both at day 7 and day 28 in the feces, although none of them was significant at term equivalent age. Interestingly, at term equivalent age several pathways related to fatty acid metabolism and fatty acid biosynthesis were significantly changed in the fecal samples, accompanied by the significant alteration of many pathways involved in amino acid metabolism. The related pathway of the urea cycle was also significantly affected at both day 28 and term equivalent age (day 28: p = 0.023, term: p = 0.001) in the feces. Tyrosine metabolism, connected to the synthesis of the neurotransmitter dopamine, was significantly altered at day 28 and term equivalent age in feces (day 28: p = 0.034, term: p = 0.005). Bile acid biosynthesis was also affected at day 7 and day 28 in feces, although it was not statistically significant (day 7: p = 0.077, day 28: p = 0.061). The metabolism of several amino acids was significantly modulated in plasma at day 3, day 7 and day 28, while no such effect was observed at term equivalent age (Fig. 2A). This includes the metabolism of glutamate, a neurotransmitter with an important role in the central nervous system, at day 3 and day 28 (day 3: p = 0.003, day 28: p = 0.007).

Also, in the plasma, pathways involved in carbohydrate metabolism were significantly altered at day 7 and to a lesser extend at day 28. Sialic acid metabolism at day 28 was significantly modulated in both plasma and feces. This includes the metabolism of the sialic acid N-acetylneuraminic acid, which participates in the brain as an integral part of ganglioside structure in synaptogenesis.^32^ Interestingly, bile acid biosynthesis was enriched in the plasma only at term equivalent age, although not considered significant (term: p = 0.059), while in the feces it played a role at day 7 and day 28. Also, biopterin metabolism was affected significantly in the plasma at term equivalent age, while it was significant in the feces at each sampling time point. In general, a development over time is apparent between different sampling time points in both, plasma and feces. For example, in the feces at day 7, carbohydrate metabolism dominated, at term equivalent age amino acid metabolism and at day 28 a mix of both was observed closing the gap between the former and later time point. Also, in the plasma a similar trend as seen in the PCA (Fig. 1B) can be observed, where the overall differences between samples is smaller at term equivalent age. In Fig. 2C the intersection of altered metabolic pathways between plasma and feces can be seen. In general, differences in abundance of potentially relevant metabolites between the groups were already observed in the early time points. In Fig. 2A, metabolites with altered levels between the experimental groups at their respective sampling time points are highlighted. Overall, several metabolites with high biological activity and/or neuroactivity were significantly different between control and pathological group at opposed sampling points.

### 2.2 Neuroactive metabolites are altered in feces of infants with short term pathological outcome/brain injury seen at term age equivalent

Several compounds with the potential to impact the central nervous system were detected in fecal samples, exhibiting significantly altered abundance between the groups (Fig. 3A). Indoxyl sulfate was found on day 7 in the majority of samples from the pathological group while it was not observed in the control group. Indoxyl sulfate is an indole derivative and a product of tryptophan metabolism. Particularly high levels were found in urine of children with autism spectrum disorder (ASD).^11^ An important property of indoxyl sulfate is its uremic neurotoxicity, indicative of chronic kidney disease, but also neurodegenerative disorders, such as Alzheimeŕs disease, Parkinsońs disease, major depressive disorder and multiple sclerosis.^33^ In contrast to the association with neurodegenerative disorders, indoxyl sulfate may also act against neuroinflammation, as an agonist of the aryl hydrogen receptor (AhR).^34^ Histamine was elevated in the pathological group at day 7 exhibiting a fold change log_2_ of 6.9, but a p-value above the significance threshold. It is an important neurotransmitter within the human body, especially in relation to immune cells. However, it has been established that bacteria present in the human gut microbiome can secrete histamine and thus influence the signaling pathways in humans, especially in asthma patients.^35^ In addition, histamine influences various brain functions such as control of pituitary hormone secretion, suppression of eating, and cognitive functions.^36^ Another compound found significantly modulated at day 7 is indolelactic acid (FC log_2_ = 4.9, p = 0.048), a metabolite produced by *Bifidobacterium*^37^, which was found to impact immune function in early life^12, 38^ and to be elevated in children with ASD.^11^ Moreover, it has also been suggested to exhibit neuroprotective properties in infants.^12^ It is closely associated with severe diseases, such as acute myocardial ischemia, and its levels were found to correlate strongly with the inflammation response in humans.^39^ Another metabolite with altered abundance at day 7 was tryptamine, which is one of the bacterial neuroactive metabolites generated through one of the TRYP6 pathways starting from the amino acid tryptophan.^10^ It has been associated with neurotoxicity and neurodegeneration, such as in case of Alzheimeŕs disease.^40–43^ Tryptamine is produced by multiple bacterial taxa including *Holdemania, Desulfovibrio, Yersinia, Tyzzerella, Bacillus, Clostridium* and *Ruminococcus* and to a lesser extent *Burkholderia* and *Pseudomonas* and has been implicated in the onset of ASD and schizophrenia.^8^ At day 7 in feces, a fold change log_2_ above 3 was observed for tryptamine but was not deemed to be significant (p = 0.269). The same happened in the case of histamine and cytosine, where a small number of missing values had a great impact on significance. 5-Hydroxyindole was found decreased in the pathological group at day 28 (FC log_2_ = -2.7, p = 0.005). It can affect the alpha-7 nicotinic receptor (α7nAChR),^44–46^ which has a potential role in the reduction of inflammatory neurotoxicity in sepsis, myocardial infarction, stroke, and Alzheimer’s disease.^47–49^ One interesting finding at day 7 was the increase of the steroid sulfates epipregnenolone sulfate (FC log_2_ = 2.7, p = 0.004), pregnenolone sulfate (FC log_2_ = 1.8, p = 0.109), corticosterone sulfate (FC log_2_ = 1.4, p = 0.048), 3β,16α- dihydroxyandrostenone sulfate (FC log_2_ = 1.0, p = 0.073) and 16α-Hydroxy DHEA 3-sulfate (FC log_2_ = 1.2, p = 0.073), in the pathological group. Epipregnenolone sulfate and pregnenolone sulfate are neurosteroids and agonists of TRPM3^50^ and negative allosteric modulators of GABA_A_^51^ and N-methyl-D-aspartate (NMDA) receptors^52^. Hypofunction of NMDA receptors was associated with impairment of synaptic plasticity.^53^ Steroids play a role in neuro modulation and as a contributor to gut-brain axis signaling^54^ and the sulfonation of steroids by gut bacteria was associated with the alteration of immune cell trafficking in the host.^55^

**Fig. 3.**
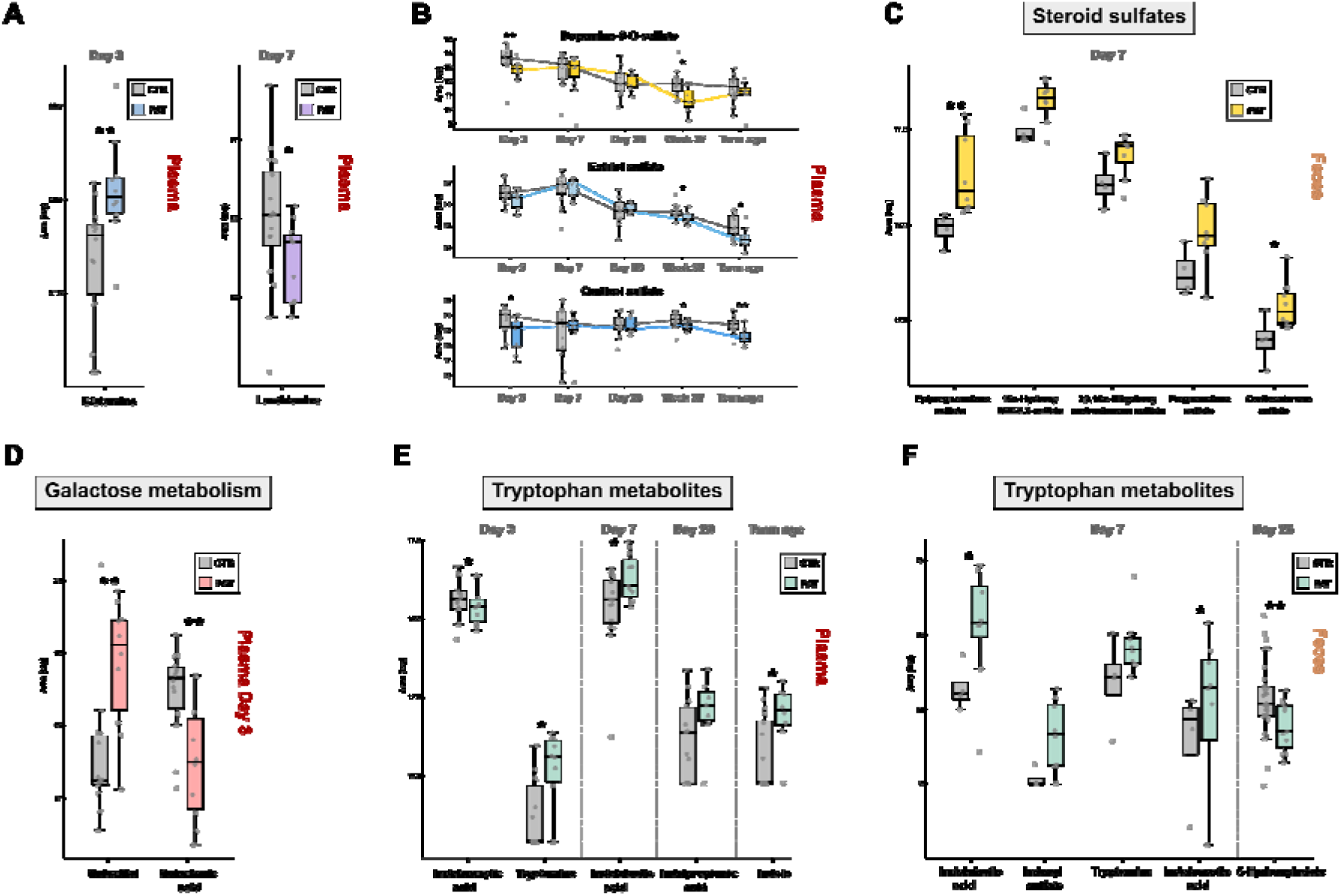
(A) Boxplots of glutamine and lanthionine in plasma. (B) Boxplots over the sampling time points of dopamine-3-O-sulfate and the steroid sulfates estriol sulfate and cortisol sulfate in plasma. (C) Boxplots of elevated steroid sulfates in the pathological group in feces at day 7. (D) Boxplots of the galactose metabolites galactitol and galactonic acid at day 3 in plasma. (E/F) Boxplots of several tryptophan metabolites with altered abundance between experimental groups in plasma and feces.

### 2.3 Metabolic profiles in plasma are already differentiated three days after delivery

Several amino acids were significantly increased in plasma samples from the pathological group at day 3, mainly Ala, Asp, Ser, Gly, Pro, Lys, homoserine, citrulline and glutamine (Tab.1), which is the principal excitatory neurotransmitter in the brain.^56^ This observation is in accordance with the results from the pathway enrichment analysis (section 2.2) (Fig. 2A, 2B), in which the metabolism of Ala, Asp, Ser, Gly and Glu was significantly affected. Another elevated metabolite at day 3 was a sugar alcohol which corresponds to galactitol or one of its stereoisomers mannitol and sorbitol (p = 0.005, FC log_2_ = 1.3) (Fig. 3D). Galactitol is a reduction product of galactose and highly elevated levels are associated to the presence of galactosemia^57^ and other genetic diseases marked by the deficiency in galactitol metabolism. In addition, it has been implicated for its potentially harmful neurotoxic properties at elevated concentrations and is connected to brain damage^58, 59^ and retinopathy.^57, 60^ Sorbitol, a reduction product of glucose, was associated to peripheral neuropathy^61^ and also retinopathy^60^ like galactitol. Mannitol, an commonly used additive in medication, elevates blood plasma osmolality, thereby reducing cerebral edema, elevated intracranial pressure, and cerebrospinal fluid volume and pressure.^62^ Although a reference standard for galactitol was acquired (which confirms the identity at an identification confidence level 1, SI Fig. S5)^63^ it cannot be excluded that it is its stereoisomere mannitol/sorbitol or a mixture of them, since their retention time and MS/MS spectra might be identical. Interestingly at the same time, a decrease in the alternative galactose metabolite galactonic acid (day 3: p = 0.007, FC log_2_ = -1.1) was observed (Fig 3D) compared to the control group, indicating that there might be an altered metabolism of galactose in the experimental group shortly after birth. Tryptamine was also found increased in the pathological group at day 3 (p = 0.011, FC log_2_ = 1.7) and decreased at day 7 (p = 0.048, FC log_2_ = -1.4) (Fig. 3E). At day 28, elevated levels of *p*-cresol sulfate were observed in the pathological group (p = 0.067, FC log_2_ = 1.1) which is a neurotoxic metabolite of bacteria-derived *p* cresol. Bacteria can produce cresol from the Tyr present in the gut, whereas liver metabolism results in p-cresol sulfate production.^64^ *p*-cresol was associated with ASD and can even exacerbate such behaviour in a dose-dependent manner.^65, 66^ Apart from neurological impairments in relation to kidney disease, *p*-cresol sulfate has been associated with disease progression in Parkinsońs disease.^33, 67^ Indole was also increased in the pathological group (p = 0.023, FC log_2_ = 1.2), which together with tryptamine is synthezised via the TRYP6 pathway by *Burkholderia* and *Pseudomonas,* in addition to *Corynebacterium* and has been found in high levels in children suffering from autism spectrum disorder.^8^ In the plasma of the pathological group a decrease of lanthionine (day 7: p = 0.039, FC log2 = -1.1)(Fig. 3A), a biomarker for H_2_S production, was observed at day 7, indicating that there might be a difference in hydrogen sulfide related metabolism in the first week after birth. We detected two forms of biliverdin (biliverdin IXβ: p = 0.027, FC log_2_ = -1.4; biliverdin IXα: p = 0.060, FC log_2_ = -1.0) and two forms of bilirubin (bilirubin IXα: p = 0.038, FC log_2_ = -1.6; bilirubin IXβ: p = 0.052, FC log_2_ = -1.5), which were elevated in the control group at term equivalent age (Fig. 2D). Biliverdin is the product of heme in heme catabolism, and is further metabolized to bilirubin, which has been associated with neurological dysfunction and neurotoxicity.^68, 69^ Interestingly the same trend was also observed another product of heme 5-Oxo-δ-bilirubin (p = 0.013, FC log_2_ = -1.4), at term equivalent age in the plasma (Fig. 2D). The related metabolic pathway porphyrin metabolism was significantly affected at term equivalent age in the enrichment analysis (Fig. 2B) in section 2.2. Increased levels of lactate and ratio of lactate/pyruvate at birth has been used as an predictor of occurrence of neonatal encephalopathy^70^ in term and preterm infants. We found a similar distribution of the lactate/pyruvate ratio (SI Fig. 6) deemed a predictor for brain injury in the pathological group, although not statistically significant. In contrast we did not observe an increased level in lactate and found significantly decreased levels of pyruvate in the pathological group.

### 2.4 Correlations between gut microbiome and neuroactive compounds observed in feces

The results of the 16S rRNA gene amplicon sequencing of the fecal samples from this cohort of extremely premature infants, obtained in our previous study^2^, was correlated with the data from the targeted and untargeted metabolomics experiments of the feces (Fig. 4B) over all sampling time points and samples available for this study. *Klebsiella* overgrowth was previously associated with brain injury in extremely premature infants and could be a potential predictor for pathological neurophysiological development. We observed significant negative correlations between *Klebsiella* and hydroxyproline, phenylalanine and 5-hydroxyindole. Significant negative correlations were also observed with the neurotransmitter GABA and N-acetyl-L-aspartic acid, the precursor of the neuronal dipeptide N-acetylaspartylglutamate. Weak positive correlations were found for methyluridine, alpha-ketoglutarate and the sialic acid deamino-neuraminic acid. *Staphylococcus* exhibited strong positive correlations with trimethylaminoxide, glutamine, asparagine and the sialic acids deamino-neuraminic acid and N-acetylneuraminic acid, but was negatively correlated with the neuroactiveTRPmetabolite indole. Tryptamine, also a neuroactive TRP metabolite, was positively correlated with *Enterococcus*, as were carnitine and N-acetyl-L-aspartic acid. There was a strong negative correlation between *Enterococcus* and the abundance of the amino acid tyrosine in the feces. *Bifidobacterium* showed significant positive correlations with the TRP metabolites indolelactic acid and indoleacetic acid, GABA and the nucleobases cytosine, uracil, adenine. Negative correlations were found for the taurine conjugate CDCA. The strongest correlations between metabolites and the 16S rRNA gene amplicon data was observed in *Escherichia-Shigella*, consisting of strong negative correlations with the amino acids asparagine, arginine, glutamate, glutamine. *Escherichia-Shigella* also exhibited significant positive correlations with GABA and the TRP metabolites indole-3-carbaldehyd and indole. Several steroid sulfates, found to be differently abundant between experimental groups, were significantly negative correlated with the growth factor VEGF-A, which was associated with pathological brain development in premature infants^2^. The sialic acids and intestinal mucus components deamino-neuraminic acid, N-acetylneuraminic acid and N-Acetylgalactosamine 6-sulfate were all significantly negative correlated with IL-17a and *Escherichia-Shigella*, while being positively correlated with IL-22. The cytokine IL-17a was found increased and IL-22 decreased in premature infants with brain injury^2^.

**Fig. 4.**
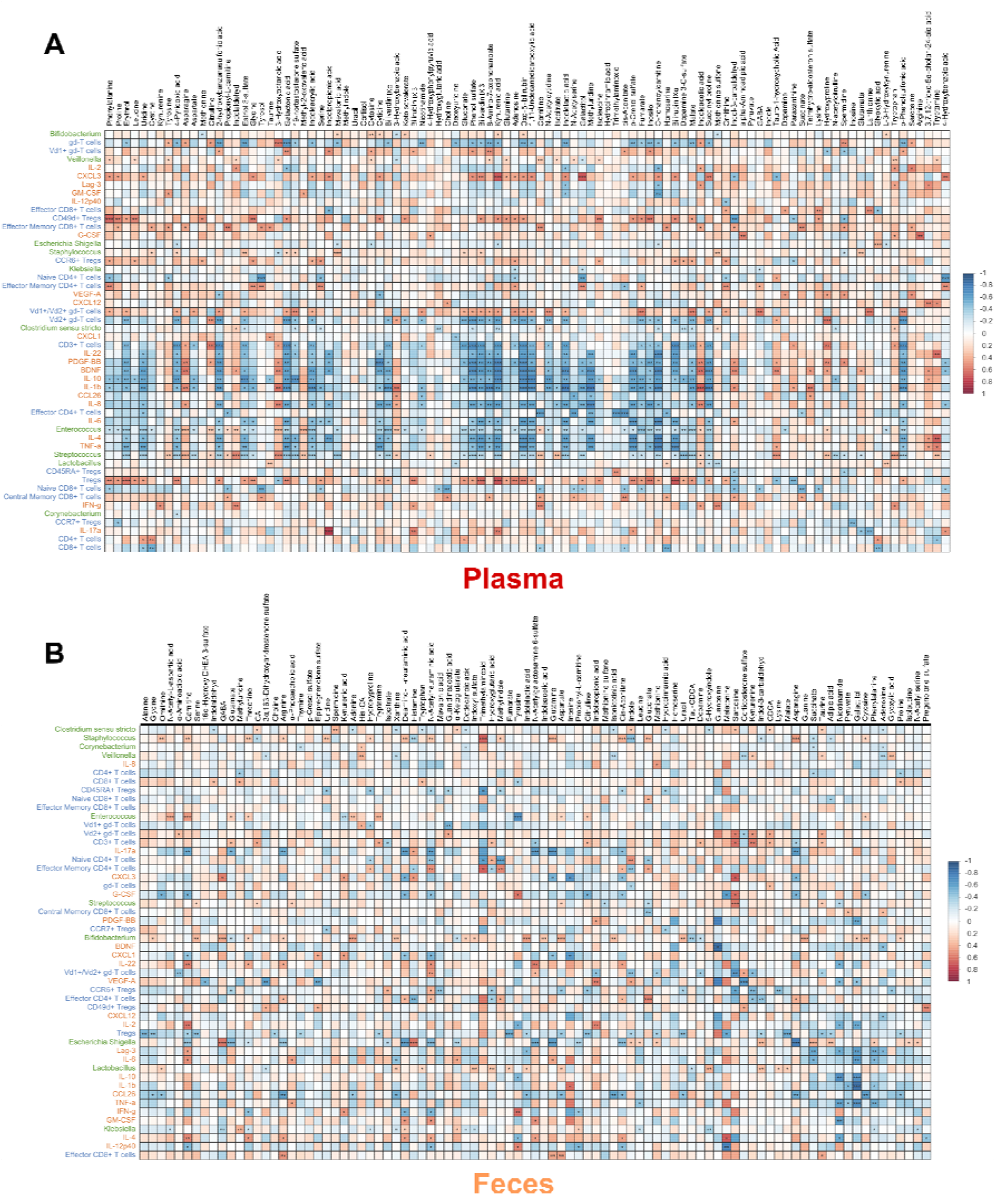
(A) Correlation matrix of 16S rRNA microbiome, T-cell, cytokine, growth factor data with metabolomics data over the time points day 3, day 7, day 28, week 32 and term equivalent age of plasma samples (T-cell: n = 27; cytokines&growth factors: n = 23, 16S rRNA microbiome: n = 78; p value: * < 0.05, ** < 0.01, *** < 0.001). (B) Correlation matrix of 16S rRNA microbiome, T-cell, cytokine, growth factor data and metabolomics data over the time points day 7, day 28 and term

In plasma (Fig. 4A) tryptamine was significantly positively correlated with pro-inflammatory factors IL-22, TNF-a and CXCL 12, which play a role in neurogenesis and neuroinflammation.^71^ Other TRP metabolites, indolpropionic acid, indolacrylic acid and indolelactic acid were negatively correlated with *Enterococcus* and *Streptococcus*, and positively with the growth factors BDNF and PDGF-BB and CXCL3, a chemokine associated with neuroprotective and anti-inflammatory activities,^72^ while for indolacetic acid and indol-3-carbaledhyd the exact opposite trend was visible. Bilirubin, biliverdin, oxo-δ-bilirubin and the compounds p-cresol sulfate and kynurenic acid, galactonic acid were all negatively correlated with the pro-inflammatory factors gd-T cells, Vd2+ gd-T cells, TNF-a, IL-6, IL-1b, IL-8, while at the same time positively correlated with the anti-inflammatory factors Tregs and CXCL3. *Enterococcus*, *Streptococcus* and gd-T cells were positively correlated with3-Hydroxyoctanoic acid as were the growth factors BDNF and PDGF-BB. For the compounds phenylalanine, tyrosine, tyrosol, serine, adenosine, galactitol, methionine sulfone and 4-hydroxybenzoic acid opposite correlation with naïve CD4+ T cells and effector memory CD4+ T cells was observable, indicating a potential effect on CD4+ T cells.

### 2.5 Bile acid profiles of plasma and feces in extremely premature infants

Recently, formerly unknown bile acid conjugates of the primary bile acid cholic acid connected to the amino acids tyrosine, phenylalanine and leucine produced by gut bacteria were discovered and were found to be increased in patients with inflammatory bowel disease.^73^ Moreover, another study reported the discovery of 145 so far undescribed bile acid conjugates^74^ by synthesizing the suspected molecules and comparing their experimental MS/MS spectra to spectral data bases. Those conjugates have been connected to disease phenotypes in Crohn’s disease, ulcerative colitis and to disease severity in IBD. In addition, the group also tested 202 isolated bacterial strains for their bile acid conjugation capabilities and found interactions between the pregnane X receptor (PXR) and some of those new bile acid conjugates. Initiated by this new discovery, we batch-synthesized more than 120 bile acid conjugates by coupling the primary bile acids cholic acid (CA), chenodeoxycholic acid (CDCA) and the secondary bile acids ursudeoxycholic acid (UDCA), deoxycholic acid (DCA), hyodeoxycholic acid (HDCA) and lithocholic acid (LCA) to 19 amino acids and taurine. A targeted LC-MS/MS method was developed and optimized for the detection and quantitation of those bile acids and their conjugates in fecal and plasma samples, including also the successful differentiation of isomeric bile acids. In the fecal samples of extremely premature infants, we identified 40 of the recently discovered bile acid conjugates of five bile acids (Fig. 5B), with the exception of lithocholic acid conjugates. With our targeted LC-MS/MS method we were also able to detect and separate two signals for the ornithine and lysine conjugates with their respective bile acids. Due to the fact that both have two amine groups it is likely that those two signals are α and ε linked versions of the bile acid conjugate. Interestingly, in the fecal samples we detected two respective version of Lys-CA and Lys- CDCA, while for ornithine-coupled bile acids only one signal was observed. This observation suggests the capability of the gut microbiome to create both α and ε linked versions of Lys-CA and Lys-CDCA. In general, the occurrence of the conjugates seems to be a highly individual factor, with some conjugates found very rarely and, in some cases, only in single samples (Fig. 5A). There seemed to be a difference in the occurrence of bile acid conjugates between the control and pathological group at day 7, especially regarding cholic acid conjugates (e.g. Phe-CA, Gln-CA), Tau-HDCA and Lys1-CDCA. Since the sample size of the control group at day 7 was limited, the observed difference might be produced by chance. In general, there was no clear distinct trend between experimental groups regarding the recently discovered bile acid conjugates. On the other hand, a clear trend between the abundance of the primary bile acids chenodeoxy cholic acid (CDCA: p = 0.033, FC log_2_ = 3.3), cholic acid p = 0.110, FC log2 = 1.6) and their taurine conjugates (Tau-CDCA: p = 0.067, FC log_2_ = -3.0; Tau-CA: p = 0.110, FC log2 = -3.0) between the experimental groups was observed (Fig. 5E). Six of the new bile acid conjugates were detected (Fig. 5D) in the plasma samples, interestingly including Orn-LCA which was not detected a single time in its free form or conjugated form in the fecal samples. Furthermore, taurine conjugates of the secondary bile acids HDCA and DCA were already observed in most samples at day 3. In addition, lower levels of Tau-CA (p = 0.036, FC log_2_ = -1.5) and Gly-CA (p = 0.110, FC log_2_ = - 1.1) in the pathological group were detected at day 3.

**Fig. 5.**
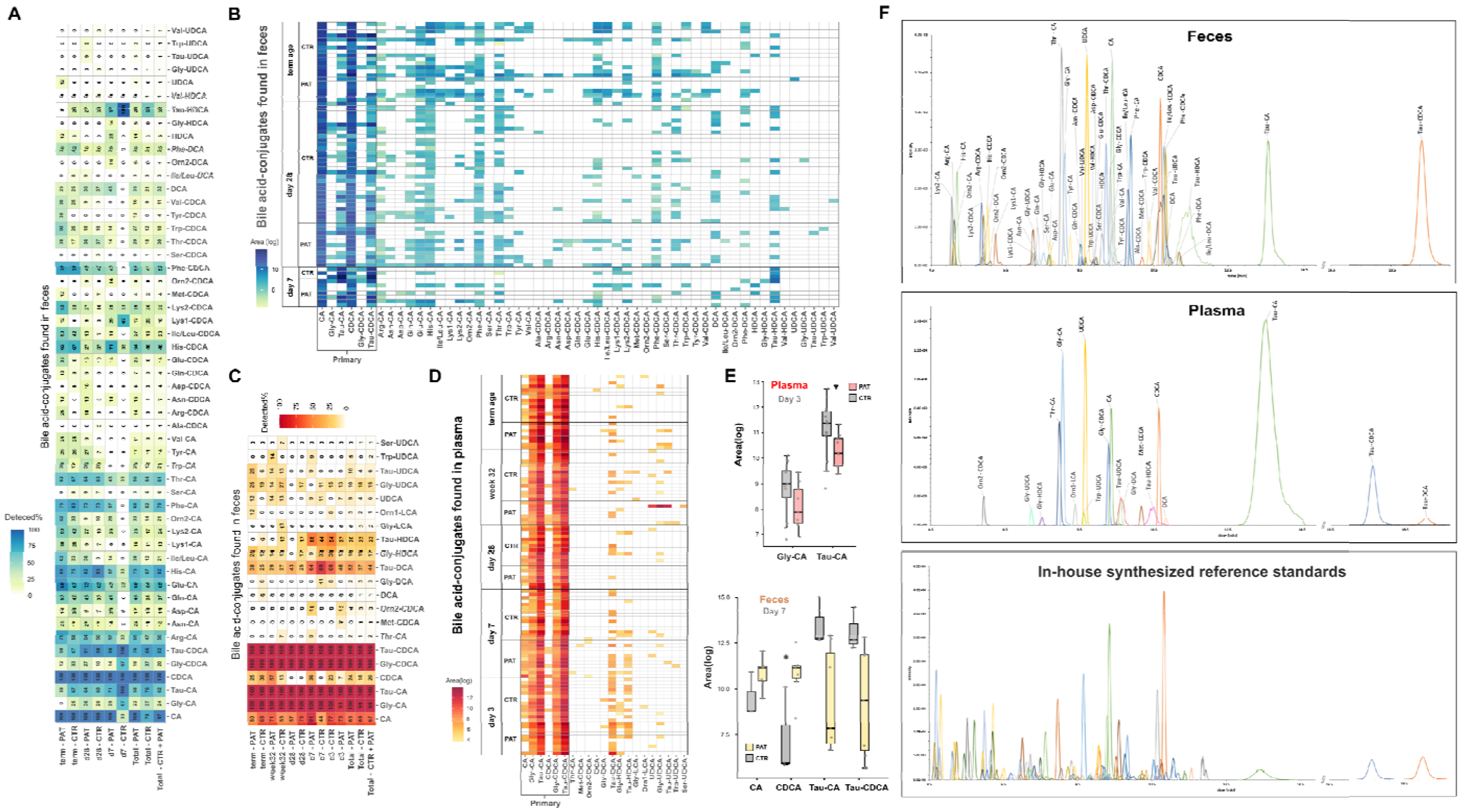
Bile acid and bile acid conjugate profiles in the feces and plasma samples obtained from extremely premature infants. Overview of single bile acid/conjugates detected (%) in the experimental groups and at different sampling time points (**A** fecal samples, **D** plasma samples). **C** Peaks of detected analytes. Bile acid profiles of individual samples (B fecal samples, **E** plasma samples). **F** Boxplots of altered bile acid/conjugates between experimental groups in plasma and fecal samples (Plasma: Tau-CA: p = 0.036, FC log_2_ = -1.6; Gly-CA: p = 0.110, FC log_2_ = -1.2; Feces: CDCA: p = 0.033, FC log_2_ = 3.3; Tau-CDCA: p = 0.067, FC log_2_ = -3.0, Tau-CA: p = 0.110, FC log_2_ = -3.0, CA: p = 0.110, FC log_2_ = 1.6).

## 3 Discussion

In this study we investigated the metabolic profiles of plasma and feces samples of extremely premature infants in two patient groups, one with age adequate findings and mild brain injuries found on MRI at term-equivalent age (CTR) and another with pathological and severe injuries (PAT) at different sampling time points during their early development. We found greater variability in the metabolic profiles of plasma and feces during the first week of life than at later time points. The general differences in the fecal and plasma metabolome varied more between sampling time points than between the two experimental groups, and were generally dominated by individual factors. Despite this expected finding, the control and pathological group clearly differed in the presence and abundance of several metabolites, some of them associated with neuroactivity. Several differences between groups were observed both in plasma and fecal samples at different time points. The beginning of those changes in neuroactive metabolites between groups was already observed at day 3 of life or in the first week after delivery, suggesting a very early onset of metabolome differentiation between experimental groups. The plasma from the pathological group exhibited elevated levels of amino acids and the neuroactive TRP metabolite tryptamine already at day 3. At day 28, a slight increase in the neuroactive and gut microbiome related compounds indole propionic acid and p-cresol sulfate in the pathological group was observed. The same trend was seen for the neurotransmitter dopamine. Plasma levels of bilirubin and biliverdin were decreased in the pathological group at term equivalent age and positive correlations with anti-inflammatory and neuroprotective factors were observed, while correlations with pro-inflammatory factors were negative. In the fecal samples the greatest differences between experimental groups were detected at day 7, including higher levels of the neuroactive TRP metabolites tryptamine, indoxyl sulfate and indolelactic acid in the pathological group. In addition, a higher abundance of the neurosteroids including epipregnenolone sulfate and pregnenolone sulfate were found in the pathological group. Overall, an increase of gut microbiome-derived metabolites, some with neuroactive potential, were found in the pathological group both in the feces and the plasma. We synthesized 120 reference standards including the majority of the newly described, bacterial-derived conjugates of bile acids and amino acids,^74^ consisting of CA, CDCA, DCA, HDCA, UDCA and LCA coupled to 19 amino acids and taurine. A targeted LC-MS/MS method for the free bile acids and their respective conjugates was developed, allowing the quantification of > 120 analytes, while ensuring the chromatographic separation of the isomeric bile acids CDCA, HDCA, DCA, UCA and their respective conjugates, through careful optimization of chromatographic separation. Utilizing this new assay, we detected 40 of the recently discovered bile acid conjugates in the fecal samples (Fig. 4A, 4B), consisting mainly of primary bile acid amino acid conjugates of CA and CDCA, but also secondary bile acid amino acid conjugates of DCA, HDCA and UDCA were detected in some samples. In the fecal sample at day 7 (Fig. 4E) a significantly higher abundance of CDCA was observed for the pathological group, while for the taurine conjugated form of CDCA an opposite trend was observed. Chenodeoxycholic acid is conjugated to taurine in the liver and the resulting Tau- CDCA is excreted via the bile into the intestinal tract where it can be reabsorbed into the blood stream (i.e., enterohepatic circulation). It can also be deconjugated back to CDCA, metabolized to secondary bile acids or other bile acid metabolites by the gut microbiome. The increase in CDCA coupled with a decrease in Tau-CDCA in the fecal samples might be an indicator for a general or a bile acid specific elevation of bacterial metabolism in the intestinal tract. In the plasma no significant changes between experimental groups in the levels of CDCA and Tau-CDCA at day 7 were found. CDCA has been shown to decrease trans- epithelial electrical resistance, increase intestinal permeability and elevate levels of IL-8, IL-6, TNF-α, and vascular endothelial growth factor release.^75^ The secondary bile acid DCA, which has the potential to increase intestinal permeability by disruption of the epithelial barrier,^76^ and its conjugates were only found in the pathological group during day 7 in the fecal sample (although not statistically significant due to the small sample size). In contrast to the fecal samples, only six of the recently described bile acid amino acid conjugates were sparingly detected in some plasma samples (Fig. 4C, 4D). It is not clear if this observation was caused by the absence of those compounds or by the expected lower concentration in the blood compared to feces, rendering them to be under the limit of detection. In general, the bile acid profile of the plasma was mainly characterized by glycine and taurine conjugated forms of the two primary bile acids CA and CDCA and the free form of CA, while the free form of CDCA was observed to a lesser degree. The taurine and glycine conjugates of the gut microbiome derived secondary bile acids HDCA, DCA and also partially UDCA were already detected in plasma samples at day 3. This observation shows that after three days post-delivery there was already a metabolically active gut microbiome established, producing several bile acid transformation products which are reabsorbed into the blood stream with the potential of affecting the host. Interestingly, the occurrence of the conjugate Tau-HDCA peaked at day 3 and day 7, was reduced at later time points, while their unconjugated forms were not observed at all. A similar trend was observed in the fecal samples, indicating there might be a time dependent shift in the metabolic capacity of the gut microbiome to produce these secondary bile acids. At day 3, higher plasma levels of the primary bile acid conjugates Tau-CA and Gly-CA were observed in the pathological group. Those changes are in line with the results of the untargeted experiments, where changes between the control and pathological group were already observed in the plasma at day 3. In general, the reported changes in bile acids at the early sampling time points in both plasma and fecal samples clearly strengthen the observations from the untargeted metabolomics experiments, where the same early onset of differentiation in metabolites between experimental groups was observed.

## 4 Limitations of the Study

Due to the nature and circumstances of extremely premature infants, it is exceptionally difficult to obtain samples in greater number from the same cohort or same patient. This results in typically smaller sample size, but also to a limited amount of possible sampling time points with greater time gaps in between. Furthermore, this results in a longitudinal sample set, in which for most patients, samples are missing at several sampling time points, making longitudinal investigations more difficult up to impossible. But not only the sample number but also the sample amount available from extremely premature infants is limited, making the sample preparation and following measurements more challenging.

Another problematic factor is the potential for different onset times in the pathological processes behind brain injury among patients. This potentially delayed onset combined with the greater time gaps between sampling time points poses as a major obstacle for a quantitative time-course investigation, especially in regard to statistical analysis, since not every patient might be in the exact same step of pathological progression at the sampling time point. At the early time points this can lead to missing values of potential key metabolites as it might be the case for tryptamine, histamine and indoxyl sulfate. For those compounds a clear trend between the groups was observed in the beginning, with the exception of a few samples where those compounds were not detected at all, which might be the case if the sample would have been collected a bit later. Missing values and missing value imputation distort the statistical analysis in those cases and makes it difficult to determine the significance of the differential metabolite abundance between groups. Those missing values combined with the small sample size increase the difficulty of analysis and reduce the statistical power and therefore the potential for deeming changes as significant. In addition, for one sampling time point (feces day 7) only a very low number of samples in the control group was available, therefore the results regarding this group have to be interpreted with caution. Despite these difficulties, we were able to collect the most comprehensive longitudinal sample set of extremely premature infants to date and obtained comprehensive and highly relevant insights by employing highly optimized sample preparation procedures and MS workflows to deal with the very low sample amounts available.

## 5 Conclusion and Outlook

The occurrence of extremely premature births is rising globally, leading to an ever-growing size of this extremely vulnerable patient group. Although survival rates of extremely premature infants have been increased by improved neonatal intensive care, a considerable number of infants suffer from severe morbidity and life-long neurodevelopmental impairment. In this work we determined the metabolic profile of plasma and fecal samples from different sampling time points in two groups of patients based on MRI findings at term equivalent age. Despite the limited sample amount and the difficult nature of the biological background of this patient group, we observed highly relevant molecular markers, indicating an early onset of differentiation between the group with brain abnormalities and the control group without any abnormalities. This distinction is characterized by a difference regarding the abundance and presence of primary bile acids/ conjugates and other compounds with potential neuroactive effects, some of them derived from the gut microbiome. Furthermore several compounds correlated to pro and anti-inflammatory factors were significantly altered between the pathological group and control group. Those compounds might be beneficial in early diagnosis of this pathological development. Furthermore, microbially derived secondary bile acids and bile acid amino acid conjugates were already present in the plasma three days after delivery, displaying the metabolic activity of the already established gut microbiome and the absorption of its metabolites into the blood stream. This indicates the potential early influence of the gut microbiome on its host is not limited to the gut-brain axis, but already takes place via direct uptake of metabolites. The findings in this study help to better understand the cause and development of brain impairments in extremely premature infants and its connection to the gut microbiome.

## 6 Material and Methods

### 6.1 Chemicals and reagents

Acetonitrile (ACN), methanol (MeOH) and water were LC-MS grade and purchased from VWR Chemicals (water) or Honeywell Riedel de-Haën (ACN, MeOH). Formic acid (FA) used as an additive for eluents was purchased from Promochem. Chemical reference standards used during LC-MS measurements were acquired from Sigma Aldrich, Carbosynth or Toronto Research Chemicals. The bile acid conjugate reference standards were synthezised as described in the supporting information (SI 1.1), and the used reagents were purchased from Sigma Aldrich. Isotopically labeled compounds used as internal standards, were purchased from Cambridge Isotope Laboratories.

### 6.2 Cohort of extremely premature infants and sample collection

In a single cohort of sixty extremely premature infants, neurophysiological development was monitored during hospitalization and brain injuries were identified by a combination of ultrasound imaging at multiple time points and magnetic resonance imaging (MRI) at term-equivalent age. Out of the cohort of 60 infants, 53 survived and 38 of the remaining infants displayed age-adequate MRIs or only mild brain injuries while 15 infants were diagnosed with severe pathological brain injuries. Out of those surviving infants, samples from 51 infants were available and used for this study (Fig. 1A). There was an almost equal gender distribution with 63% females in the control group and 53% females in the pathological group. Among the observed injuries were periventricular hemorrhagic infarctions, intraventricular hemorrhages, cerebellar hemorrhages, and periventricular leukomalacia. A reduction of brain volume and an enlargement of the subarachnoid spaces was accompanying many of those injuries. Over the course of hospitalization and when possible, fecal samples were collected, and blood was drawn at multiple time points with a focus on day 3, day 7, day 28, correction week 32 and term equivalent age. The fecal samples from all extremely premature infants were collected from diapers via collection tubes and stored at – 80 °C until further analysis. For minimizing the stress for patients, blood samples were only taken combined with clinical routine sampling. The blood samples were transferred into Lithium Heparin S-Monovettes and the plasma was separated and stored at -80 °C until further analysis. Additional information about the cohort and MRI-results and neurophysiological assessment can be found in previous work^2^.

### 6.3 Sample preparation of plasma samples

Plasma samples obtained from premature infants with pathological damage or age adequate damage were used. The samples were stored at –80 °C prior to extraction. During the whole sample preparation, the samples were kept on ice. Homogenization by vortexing was followed by the addition of the extraction solvent (ACN/MeOH, 50:50, v:v) to the homogenized sample, resulting in a 1:4 volume ratio of sample and extraction solvent. After further vortexing and sonication in an ice bath for 15 min, the sample solutions were stored overnight at -20 C°. Afterwards the samples were centrifuged at 18,000 g and 4°C for 10 min and the supernatant was transferred to a new tube. For targeted LC-MS/MS analysis of bile acid conjugates, aliquots were taken from the solution and stored at -80 °C until further use. The remaining solution was evaporated to dryness at 4°C using a SpeedVac concentrator (Labconco). The residues were reconstituted (ACN/ water, 50:50, v:v) to one fifth of the volume prior to evaporation, vortexed and sonicated in an ice bath for 15 min. After centrifugation at 18,000g and 4°C for 10 min, the supernatants were transferred to LC vials and stored at -80°C until analysis. An internal standard (^2^H -ursudeoxycholic acid) was added to the aliquots taken for the targeted LC-MS/MS measurements to a final concentration of 2 µM, followed by a transfer of the samples to LC vials and storage at -80°C until analysis.

### 6.4 Sample preparation of fecal samples

Stool samples obtained from premature infants with pathological damage or age adequate damage were used. The samples were stored at –80 °C prior to the extraction. During the whole sample preparation the samples were kept on ice. The samples were manually homogenized using a spatula and dried at 4°C using a SpeedVac concentrator (Labconco) for 60 h followed by the addition of MeOH/ACN/H_2_O (40:40:20; v:v:v) at a ratio of 50 µL for each mg of dried sample. After vortexing and sonication on ice for 10 min, the samples were homogenized in a bead shaker (4 m/s, MP FastPrep-24 5G) for 15 s and stored overnight at - 20°C. Afterwards, the samples were centrifuged at 18000 g and 4°C for 10 min and transferred to a new tube. The solution was diluted 1:1 with ACN/H_2_O (20:80; v:v) containing internal standards (^13^C_6_ -bisphenol A, ^2^H_4_ -genisteine). After centrifugation at 18,000g and 4°C for 10 min, samples were sonicated in an ice bath for 10 min to dissolve potential precipitate and transferred to HPLC vials and stored at -80°C until analysis.

The fecal samples for the targeted LC-MS/MS analysis of bile acid conjugates were dried as described above, followed by the addition of 25 µL H_2_O for each mg of dried sample and the homogenization in a bead shaker FastPrep-24 5G (4 m/s) for 10 s. Subsequently the samples were centrifuged at 18,000 x g and 4°C for 10 min and half the volume of the supernatant was transferred to a new tube and stored for additional biochemical assays. The next step was the addition of MeOH/ACN (1:1; v:v) to a final solution of MeOH/ACN/H_2_O (40:40:20; v:v:v), following the extraction with a bead shaker (4 m/s, MP FastPrep-24 5G) for 10 s. After sonication in an ice bath for 10 min, the sample solutions were stored overnight at -20 C°. Afterwards the samples were centrifuged at 18,000 x g and 4°C for 10 min and transferred to a new tube. The remaining solution was evaporated to dryness at 4°C in a SpeedVac (Labconco) and the residues were reconstituted with (ACN/H_2_O, 50:50, v:v), containing an internal standard (^2^H_4_-ursudeoxycholic acid, 2 µM), to the same volume before evaporation, vortexed and sonicated in an ice bath for 15 min. After centrifugation at 18,000g and 4°C for 10 min, the samples were transferred to HPLC vials and stored at -80°C until analysis.

### 6.5 Untargeted metabolomics experiments

For the analysis a Vanquish UHPLC system coupled to a QExactive HF quadrupole-Orbitrap (Thermo) mass spectrometer via an ESI interface was used. Two columns, a HILIC column (SeQuant ZIC-pHILIC; 5 μm, 150 x 2.1 mm) and a RP column (Waters Acquity HSS T3; 1.8 μm, 100 x 2.1 mm) were chosen for complementary and broad coverage of analytes. In both cases a precolumn made of the respective material was used. An injection volume of 3 μL, a flow rate of 0.3 mL/min and a column compartment temperature of 40°C was selected for both columns. The HILIC chromatography utilized H_2_O and ACN with 10 mM NH_4_HCO_3_ (90:10, v:v, pH 9.2, eluent A) and ACN (eluent B) for the following gradient: 0-6 min linear decrease from 75% B to 45%B, 6-7 min 45% B to 30% B, 7-10 min constant at 30% B, 10 min until 10.1 min increase from 30% B to 75% B and maintained constant at 75% B for 5 min. The RP chromatography utilized H_2_O with 0.1% formic acid (eluent A) and MeOH with 0.1% formic acid (eluent B) for the following gradient: 0-0.5 min stay at 5% B, 0.5-11.5 min linear increase to 95% B, 11.5-15.5 hold at 95% B,15.5-15.6 decrease from 95% B to 5% B, 15.6-17 min stay at 5% B. Measurements were conducted in either full scan with polarity switching mode or in positive and negative mode with data dependent MS^2^ collection. The settings of the ESI interface were: sheath gas, 48 au; auxiliary gas, 15 au; sweep gas flow, 2 au; capillary voltage, 3.5 kV (positive), 2.8 kV (negative); capillary temperature, 300°C; auxiliary gas heater, 350°C. The scan range was from m/z 62 to 900. For full scan measurements the resolution was set to 60,000 with an automatic gain control (AGC) of 1x10^6^ and a maximum injection time of 200 ms. For generating MS^2^ spectra, data dependent acquisition in combination with an inclusion list with features of interest, based on the untargeted and targeted evaluation of the MS1 data, was used. For the data-dependent MS^2^ acquisition a resolution of 60 000 with an AGC target of 1x10^6^ and a maximum injection time of 100 ms were chosen for the full scan. The settings for the MS^2^ collection were as following: resolution 30, 000, AGC target 5x10^5^, maximum injection time 120 ms, loop count 10, isolation window 1.0 m/z, stepped collision energy 30-50 eV; minimum AGC 8x10^3^.

### 6.6 Targeted LC-MS/MS for bile acids and their conjugates

A targeted LC-MS/MS for the investigation of bile acids and their conjugates was developed employing commercially purchased or self synthezised reference standards (SI 1.1). For the targeted LC-MS analysis a Dionex Ultimate 3000 UHPLC (Thermo) system coupled to a TSQ Vantage triple quadrupole mass spectrometer (Thermo) via an ESI interface was used. An Atlantis T3 column (Waters, 3 μm, 3 mm × 150 mm), including a precolumn made of the same material, at a flow rate of 0.6 mL/min was used and maintained at 40 °C. The injection volume was 5 μL. Chromatography used H_2_O with 0.1% formic acid (eluent A) and ACN/MeOH (1:1, v:v, eluent B) with 2% H_2_O and 0.1% formic acid for the following gradient: 0–1 min at 5% B; rise to 50% B until 1.5 min; increase to 95% B until 10 min; 10–13 min at 95% B; 13-13.1 min from 95% B to 75% B and maintain until 19 min before re-equilibrating at 5% B until 21 min. Negative ESI mode was used with the following ion source parameters: Capillary temperature of 300 (°C), vaporizer temperature of 300 (°C), sheath gas pressure of 40 (Arb), aux valve flow of 10 (Arb), declustering potential of 14 (V), collision gas pressure of 1.5 (mTorr) and a spray voltage of 3000 (V). The cycle time was set to 0.6 (s), while Q1 and Q3 peak width was kept at 0.7 (FWHM).

### 6.7 QC & QA measurements for LC-MS methods

For the untargeted metabolomics measurements multiple measures to assure high quality data were taken. During the sample preparation three process blanks were processed together with the samples to investigate potential contaminations during the procedure and measured together with solvent blanks at the beginning and end of each sequence. For system suitability a mixture of reference standards (2 µM, 159 compounds, SI Tab. S1) was measured at the beginning and the end of the sequence, and for monitoring potential carry over between samples, solvent blank injections (ACN/H_2_O, v:v, 1:1) were measured. A pooled QC prepared from all samples was injected at the beginning, the end and after every five samples and analyzed throughout the sequence and were used together with internal standards (^13^C_6_ -bisphenol A, ^2^H_4_ -genisteine, 2 µM) for monitoring potential analytical deviations during the measurement. A SRM 1950 sample (metabolites in frozen plasma, NIST, Gaithersburg, USA) was extracted according to protocol^77^ and measured at the beginning and the end of a sequence.

During the targeted LC-MS/MS measurements process blanks and solvent blanks were measured at the beginning and end of each sequence. A pooled QC, spiked with the reference standards (n = 126, SI Fig. S1), was injected at the beginning, the end and after every 20 samples, and was used together with an internal standard (^2^H -ursudeoxycholic acid, 2 µM) for monitoring potential analytical deviations during the measurement. The injection order of the samples during LC-HRMS and LC-MS/MS experiments was randomized to avoid any experimental group specific bias. The NTA Study Reporting Tool (SRT)^78^ was used as guidance for the construction of the materials and methods section to ensure high reporting quality.

### 6.8 Data processing and integration of LC-MS acquired data

Thermo Fisher LC-HRMS(/MS) raw data files from the untargeted metabolomics measurements were converted into mzXML files, centroided and separated into two files according to polarity using Proteo Wizard(3.0.18)^79^. All experimental sampling time points from their respective biological matrix were processed together. Data pre-processing was done using the R (4.1) package XCMS^80^ (3.14), starting with the import of the mzXML files. The next step was peak detection using the centWave algorithm (parameters: ppm = 5, peakwidth = 5/15; snthresh = 10, prefilter = 3/5000) followed by the removing of peaks with a too wide peak width. The retention time between samples were aligned employing the obiwarp algorithm (parameters: binSize = 0.6, distFun = "cor_opt") with a previous alignment of pooled QC samples followed by the alignment of the samples. The detected peaks were grouped to features with a minimum threshold of 40% in at least one experimental group (parameters: bw = 1, minFraction = 0.4, minSamples = 5). Features missing a peak in certain samples were integrated in the respective retention time width of the feature. The resulting feature list was further processed by the R package CAMERA (1.48)^81^ (parameters: perfwhm = 0.6, ppm = 5), creating adduct and isotopologue annotations. Features annotated as isotopologues and features with an adduct annotation different from [M+H] or [M-H] were removed from the feature list. In addition, features with a signal in the process blank or solvent blank higher than 20 % of the median signal in the pooled QC were excluded. All features eluting in the void volume were also removed from the feature list (HILIC: 1 min, RP: 0.65 min). The python based package NeatMS (0.9)^82^, based on neuronal networking, was used to classify the peak quality. Features mainly consisting of the peak label “Noise”/ “Low Quality” were filtered out. For each sampling time point a subset of the feature list was created and further processed. Features with more than 20% missing values were removed and the missing values were imputed by using half of the minimum value. Fold changes (log_2_) between groups were determined for each time point, and features within a threshold of 2 < fold change log_2_ < -2 were considered for further MS2 experiments and annotation. In addition to the untargeted evaluation, a targeted evaluation of the data obtained from the untargeted experiments was done using the software Skyline (20.2)^85^, including compounds and features of high interest and/or compounds with an reference standard available. Feature annotation was carried out by in silico fragmentation employing SIRIUS 4^83^ (*parameter: 5 ppm; possible Ionizations [M-H]-, [M+Cl], [M+H]+, [M+Na]+; DB: Bio database*), based on the received score and manual reviewing, and was expanded by comparing experimental MS^2^ spectra with spectral libraries using GNPS^84^ (parameters: Score Threshold: 0.7, Precursor Ion Mass Tolerance: 0.05, Min Matched Peaks: 5) whenever possible. Features annotated by employing *in sillico* fragmentation were assigned an identification confidence level of 3, features with an unambiguous match in a spectral library with an identification confidence level of 2.a, while compounds with an identity confirmed by reference standards, were assigned an identification confidence level of 1.^63^ The raw files obtained from the LC-MS/MS experiments targeting bile acids and their conjugates were also processed and quantified by Skyline (20.2). Fold changes (log_2_) between groups were determined and corresponding p-values were calculated using Wilcoxon Rank sum test. FDR was controlled using the ‘qvalue’ (2.28)^86, 87^ R package with a cutoff of < 0.2. For a putative Gene Set Enrichment Analysis (GSEA) and over-representation analysis (ORA, mummichog^29^)) the ‘MS Peaks to Pathways’ tool from the web application MetaboAnalyst (5.0)^30^ was used (*parameters: p-value cutoff: 0.1, mummichog+GSEA, min. 5 entries, MFN DB*). For this purpose, the data sets of the HILIC and the RP chromatography and positive and negative polarity mode were combined to one data set for each time point. Multivariate statistical analysis (PCA) was done using the R package ‘MixOmics’ (6.20)^88^ on the auto-scaled and combined data of both positive and negative polarity of RP and HILIC chromatography. Correlation analysis was done using the R package ‘Hmisc’ (4.7)^89^ based on Spearman’s correlation and for the PERMANOVA and dispersion analysis the R package ‘vegan’ (2.3)^90^ was used.

## Data Availability

All data produced in the present study are available upon reasonable request to the authors

