## Supplementary Information for "Neuroactive metabolites and bile acids are altered in extremely premature infants with brain injury"

### Supporting Information

#### List of content

1 Experimental ..................... 2

1.1 Batch Synthesis of bile acid conjugates ..................... 2

Fig. S1. Overview of synthezised standards ..................... 2
Fig. S2. $MS^2$ spectra of synthezised standards and reference standards ..................... 2

1.1 LC measurement sequence ..................... 3

Fig. S3. LC injection scheme ..................... 3

1.1 Chemicals and reagents ..................... 4

Tab. S1. List of reference standards. ..................... 4

1.2 Targeted LC-MS/MS method for bile acids and bile acid conjugates ..................... 6

Fig. S4. Extracted ion chromatograms of positive identified analytes in plasma ... 6
Fig. S5. Extracted ion chromatograms of positive identified analytes in feces ...... 8

2 Additional plots and tables ..................... 12

Fig. S6. Distribution of lactate/pyruvate ration ..................... 12
Tab. S3. Overview of highlighted metabolites in plasma ..................... 13
Tab. S4. Overview of highlighted metabolites in feces ..................... 15

3 Compound identification ..................... 16

Fig. S7. Galacitol reference standard ..................... 16

### 1 Experimental

#### 1.1 Batch Synthesis of bile acid conjugates[1]

A solution of a single bile acid (40 µL, 2.5 mM, 1 eq) was combined with freshly prepared solutions of triphenylphosphine (40 µL, 10 mM, 4 eq) and dipyridyldisulfide (40 µL, 10 mM, 4 eq), vortexed and incubated for 1 min. A amino acid stock solution (5 µL, 2 mM, 0.25 eq), containing 10 amino acids, was added and the reaction mixture was incubated on a thermo-shaker (TS-100, Biosan) at 60 °C for 30 min, followed by the addition of 300 µL of ACN. Taurine conjugates were synthezised in the same manner. This procedure was repeated for all bile acids until all desired compounds were acquired. From each mixture 150 µL were taken, combined, evaporated and reconstituted in 150 µL ACN to generate a concentrated standard.

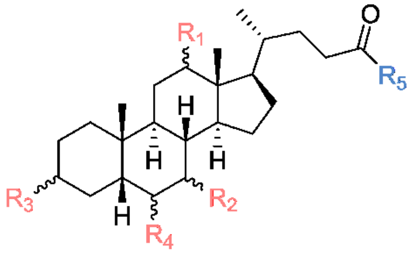

| | |
|---|---|
| $R_{1,2,3}$ = OH, $R_4$ = H | Cholic acid |
| $R_{1,3}$ = OH, $R_{2,4}$ = H | Deoxycholic acid |
| $R_{2\alpha,3}$ = OH, $R_{1,4}$ = H | Chenodeoxycholic acid |
| $R_{2\beta,3}$ = OH, $R_{1,4}$ = H | Ursodeoxycholic acid |
| $R_{3,4}$ = OH, $R_{1,2}$ = H | Hyodeoxycholic acid |
| $R_3$ = OH, $R_{1,2,4}$ = H | Lithocholic acid |

$R_5$=

| | | |
|---|---|---|
| Ala | His | Tau |
| Arg | Ile | Thr |
| Asn | Leu | Trp |
| Asp | Lys | Tyr |
| Gln | Met | Val |
| Glu | Phe | Orn |
| Gly | Ser | |

**Fig. S 1** Overview of synthesized bile acid amino acid and taurine conjugates.

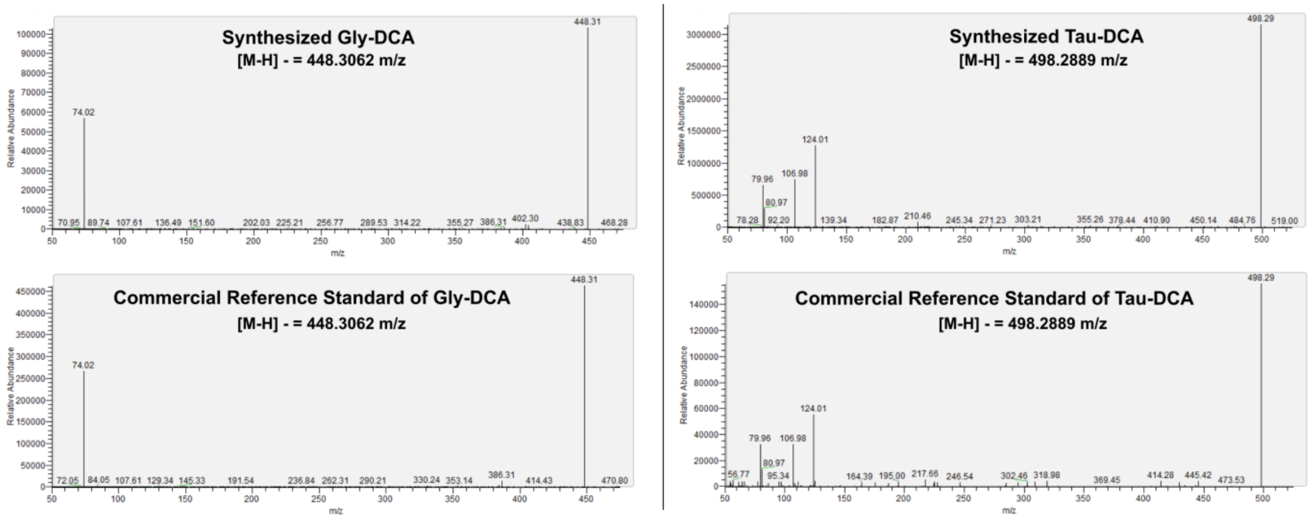

**Fig. S 2** MS$^2$ spectra of synthesized bile acid conjugates compared to commercial reference standards

[1] Matsueda et al., "Peptide Synthesis by Oxidation-Reduction Condensation. II. The Use of Disulfide as an Oxidant."

#### 1.1 LC measurement sequence

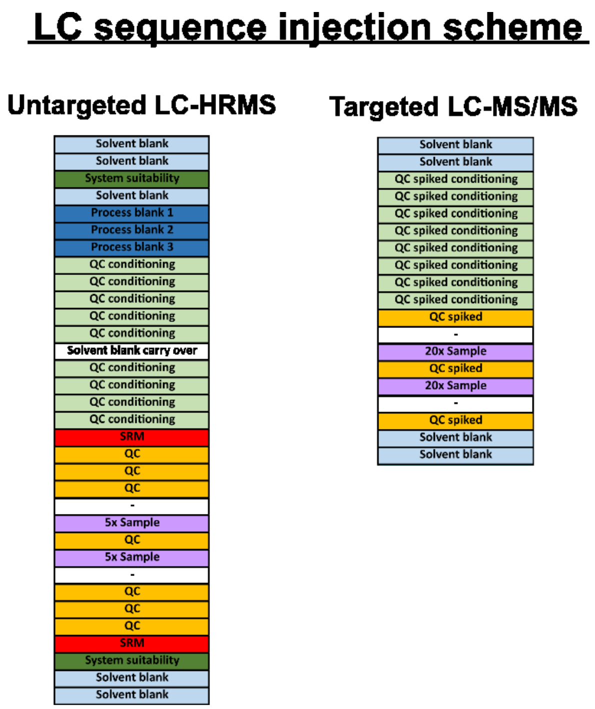

**Fig. S3.** Overview of the injection scheme of LC sequences for untargeted LC-HRMS and LC-MS/MS.

#### 1.1 Chemicals and reagents

**Tab. S1.** Overview of used reference standards for compound identification or for system suitability.

| Compound | Exact mass | CAS number | Compound | Exact mass | CAS number |
|---|---|---|---|---|---|
| 1-Methylhydantoin | 114.0429 | 616-04-6 | Indol-3-aldehyd | 145.0528 | 487-89-8 |
| 1-Methylnicotinamide | 136.0637 | 3106-60-3 | Indole | 117.0578 | 120-72-9 |
| 2-(Carbamoylamino)butanedioic acid | 176.0433 | 923-37-5 | Inosine | 268.0808 | 58-63-9 |
| 2-Deoxyadenosine 5'-monophosphate | 331.0682 | 653-63-4 | Inosine 5'-monophosphate | 348.0471 | 131-99-7 |
| 2-Deoxycytidine | 227.0906 | 951-77-9 | Inositol | 180.0634 | 551-72-4 |
| 2-Deoxycytidine 5'-Monophosphate | 307.0569 | 1032-65-1 | Isocitrate | 192.027 | 320-77-4 |
| 2'-Deoxyuridine | 228.0746 | 951-78-0 | Isoguanosine | 283.0917 | 1818-71-9 |
| 2-Phosphoglycerate | 185.9929 | 2553-59-5 | Isoleucine | 131.0946 | 443-79-8 |
| 3-Indoleacetic acid | 175.0633 | 87-51-4 | Ketoisovalerate | 116.0473 | 759-05-7 |
| 3-Indolpropionic acid | 189.079 | 830-96-6 | Kynurenic acid | 189.0426 | 492-27-3 |
| 3-Methyl-2-oxovaleric acid | 130.063 | 1460-34-0 | Kynurenine | 208.0848 | 343-65-7 |
| 3-Methylcytidine | 257.1012 | 2140-64-9 | Lactate | 90.0317 | 113-21-3 |
| 3-Methylindole | 131.0735 | 83-34-1 | L-Citrulline | 175.0957 | 372-75-8 |
| 3-Phosphoglycerate | 185.9929 | 820-11-1 | L-Cystathionine | 222.0674 | 56-88-2 |
| 4-Hydroxy-proline | 131.0582 | 6912-67-0 | Leucine | 131.0946 | 61-90-5 |
| 5'-Deoxy-5'-Methylthioadenosine | 297.0896 | 2457-80-9 | L-Ornithine | 132.0899 | 70-26-8 |
| 5-Methyluridine | 258.0852 | 1463-10-1 | Lysine | 146.1055 | 56-87-1 |
| 6-Phosphogluconate | 276.0246 | 921-62-0 | Malic acid (Malate) | 134.0215 | 320-77-4 |
| Adenine | 135.0545 | 73-24-5 | Mannitol | 182.079 | 69-65-8 |
| Adenosine | 267.0968 | 58-61-7 | Mannitol 1-phosphate | 262.0454 | 15806-48-1 |
| Adenosine 3',5'-cyclic monophosphate | 329.0525 | 60-92-4 | Mannose | 180.0634 | 530-26-7 |
| Adenosine 3'-monophosphate | 347.0631 | 84-21-9 | Melatonine | 232.1212 | 73-31-4 |
| Adenosine 5'-monophosphate | 347.0631 | 61-19-8 | Methionine | 149.051 | 63-68-3 |
| Adenosine 5'-triphosphate | 506.9957 | 56-65-5 | Methionine sulfone | 181.0409 | 7314-32-1 |
| Adenosine diphosphate | 427.0294 | 58-64-0 | Mevalonic acid | 148.0736 | 17817-88-8 |
| Alanine | 89.0477 | 302-72-7 | N4-Acetylcytidine | 285.0961 | 3768-18-1 |
| alpha-Aminoadipic acid | 161.0688 | 542-32-5 | N-Acetyl-Asp-Glu | 304.0907 | 3106-85-2 |
| alpha-Ketoglutarate | 146.0215 | 328-50-7 | N-Acetyl-L-aspartic acid | 175.0481 | 997-55-7 |
| Arginine | 174.1117 | 74-79-3 | N-Acetyl-serine | 147.0532 | 16354-58-8 |
| Argininosuccinic acid | 290.1226 | 2387-71-5 | NAD+ | 663.1091 | 53-84-9 |
| Asparagine | 132.0535 | 70-47-3 | NADH | 665.1248 | 58-68-4 |
| Aspartic acid | 133.0375 | 56-84-8 | NADP+ | 743.0755 | 604-79-5 |
| Betaine | 117.079 | 107-43-7 | NADPH | 745.0911 | 53-57-6 |
| Biotin | 244.0882 | 58-85-5 | Nicotinamide | 122.048 | 98-92-0 |
| Carnitine | 161.1052 | 406-76-8 | Octopamine | 153.079 | 104-14-3 |
| Choline | 104.1075 | 67-48-1 | Oxaloacetic acid | 132.0059 | 328-42-7 |
| cis-Aconitate | 174.0164 | 585-84-2 | Palmitic acid | 256.2402 | 57-10-3 |
| Citrate | 192.027 | 126-44-3 | p-cresol | 108.0575 | 106-44-5 |

| | | | | | |
|---|---|---|---|---|---|
| Cysteic acid | 169.0045 | 13100-82-8 | p-cresol sulfate | 188.0143 | 3233-58-7 |
| Cysteine | 121.0197 | 52-90-4 | Phenylalanine | 165.079 | 63-91-2 |
| Cysteinyl-glycine | 178.0412 | 19246-18-5 | Phosphocreatine | 211.0358 | 67-07-2 |
| Cystine | 240.0238 | 923-32-0 | Proline | 115.0633 | 4305-67-3 |
| Cytidine | 243.0855 | 65-46-3 | Propionyl-L-carnitine | 217.1314 | 17298-37-2 |
| Bilirubin IXβ | 584.2634 | 635-65-4 | Pseudouridine | 244.0695 | 1445-07-4 |
| Biliverdin IXβ | 582.2478 | 114-25-0 | Pyruvate | 88.016 | 57-60-3 |
| Cytosine | 111.0433 | 71-30-7 | Ribose | 150.0528 | 10257-32-6 |
| Deoxyguanosine triphosphate | 506.9957 | 2564-35-4 | Ribose-5-phosphate | 230.0192 | 4300-28-1 |
| Dihydroxyacetonephosphate | 169.998 | 57-04-5 | Ribulose-5-phosphate | 230.0192 | 551-85-9 |
| Dihydroxyisovalerate | 134.0579 | 19451-56-0 | S-(Adenosyl)-methionine | 398.1372 | 29908-03-0 |
| Dopamine | 153.079 | 51-61-6 | Sarcosine | 89.0477 | 107-97-1 |
| Erythrito | 122.0579 | 149-32-6 | Sedoheptulose-7-phosphate | 290.0403 | 2646-35-7 |
| Erythrose-4-phosphate | 200.0086 | 585-18-2 | Seleno-methionine | 196.9955 | 3211-76-5 |
| Flavinadenin dinucleotide | 785.1571 | 146-14-5 | Serine | 105.0426 | 56-45-1 |
| Fructose | 180.0634 | 30237-26-4 | Serotonine | 176.095 | 50-67-9 |
| Fructose-1,6-bisphosphate | 339.9961 | 34693-23-7 | Spermidine | 145.1579 | 124-20-9 |
| Fructose-6-phosphate | 260.0297 | 643-13-0 | Spermine | 202.2157 | 71-44-3 |
| Fumarate | 116.011 | 142-42-7 | Succinate | 118.0266 | 56-14-4 |
| Galactose | 180.0634 | 10257-28-0 | Taurine | 125.0147 | 107-35-7 |
| Gamma-aminobutyric acid | 103.0633 | 56-12-2 | Thiamine | 265.1123 | 67-03-8 |
| Gluconate | 196.0583 | 526-95-4 | Threonine | 119.0582 | 72-19-5 |
| Glucose | 180.0634 | 2280-44-6 | Thymidine | 242.0903 | 50-89-5 |
| Glucose-1-phosphate | 260.0297 | 59-56-3 | Thymidine 5-monophosphate | 322.0566 | 14057-65-9 |
| Glucose-6-phosphate | 260.0297 | 299-31-0 | Thymidine 5'-triphosphate | 481.9893 | 365-08-2 |
| Glutamate | 147.0532 | 56-86-0 | Thymine | 126.0429 | 65-71-4 |
| Glutamine | 146.0691 | 56-85-9 | Trehalose | 342.1162 | 99-20-7 |
| Glutamyl-cysteine | 250.0623 | 686-58-8 | Trimethylaminoxid | 75.0684 | 1184-78-7 |
| Glutathione, oxidized | 612.152 | 27025-41-8 | Tryptamine | 160.1 | 61-54-1 |
| Glutathione, reduced | 307.0838 | 70-18-8 | Tryptophan | 204.0899 | 73-22-3 |
| Glycine | 75.032 | 56-40-6 | Tyrosine | 181.0739 | 60-18-4 |
| Glyoxylic acid | 74.0004 | 298-12-4 | Uracil | 112.0273 | 66-22-8 |
| Guanidineacetic acid | 117.0538 | 352-97-6 | Uridine | 244.0695 | 58-96-8 |
| Guanine | 151.0494 | 73-40-5 | Uridine 5'-diphosphate | 404.0022 | 58-98-0 |
| Guanosine | 283.0917 | 118-00-3 | Uridine 5'-monophosphate | 324.0359 | 58-97-9 |
| Guanosine 3',5'-cyclic monophosphate | 345.0474 | 7665-99-8 | Uridine 5'-triphosphate | 483.9685 | 63-39-8 |
| Guanosine 5'-diphosphate | 443.0243 | 146-91-8 | Valine | 117.079 | 72-18-4 |
| Guanosine 5'-triphosphate | 522.9907 | 86-01-1 | Xanthine | 152.0334 | 69-89-6 |
| Guanosine-5'-monophosphate | 363.058 | 85-32-5 | Xylose | 150.0528 | 10257-31-5 |
| Histidine | 155.0695 | 71-00-1 | | | |
| Homocysteine | 135.0354 | 6027-13-0 | | | |
| Homoserine | 119.0582 | 1927-25-9 | | | |
| Hydrocinnamic acid | 150.0681 | 501-52-0 | | | |
| Hydroxyglutaric acid | 148.0372 | 2889-31-8 | | | |

#### 1.2 Targeted LC-MS/MS method for bile acids and bile acid conjugates

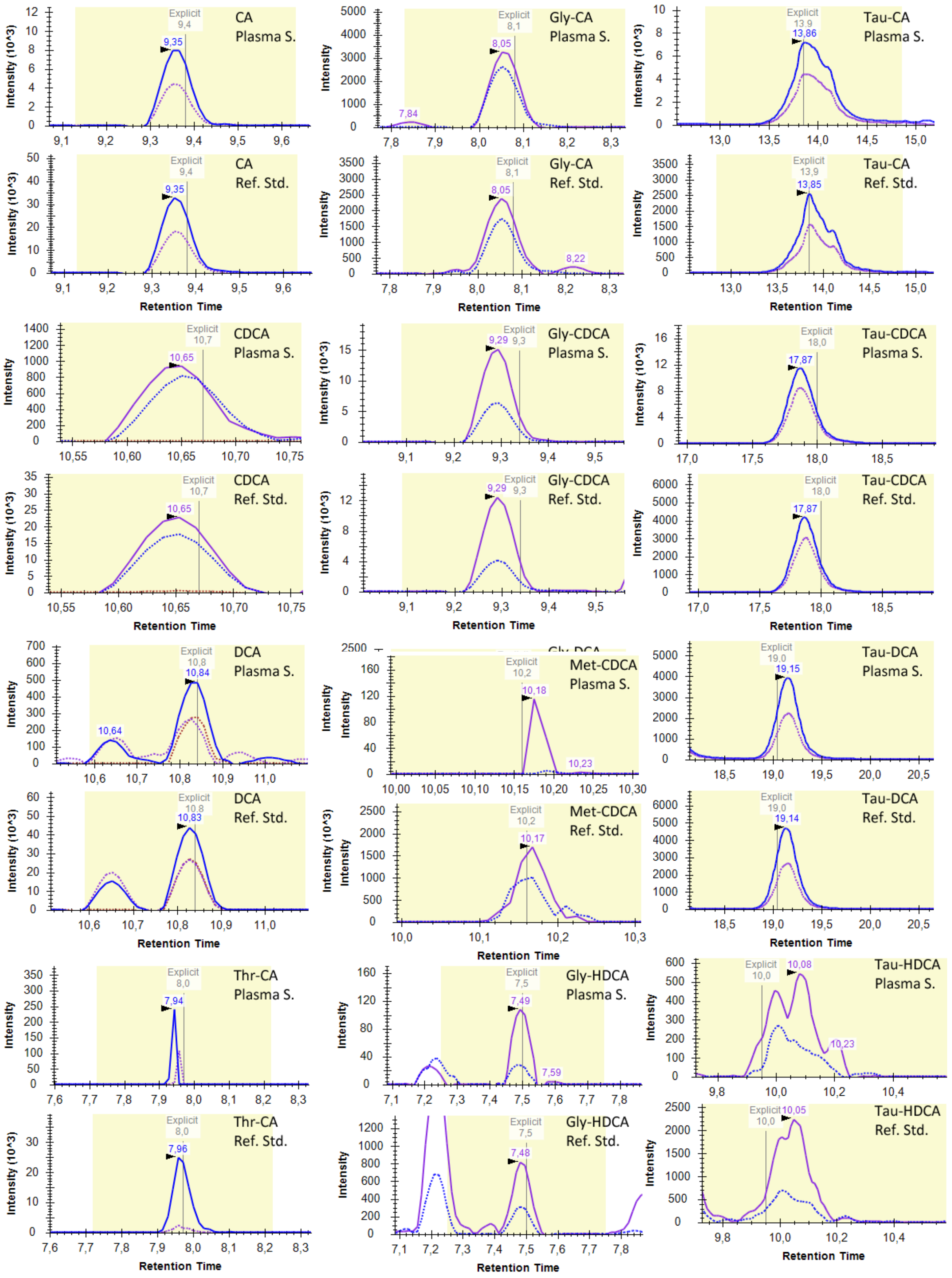

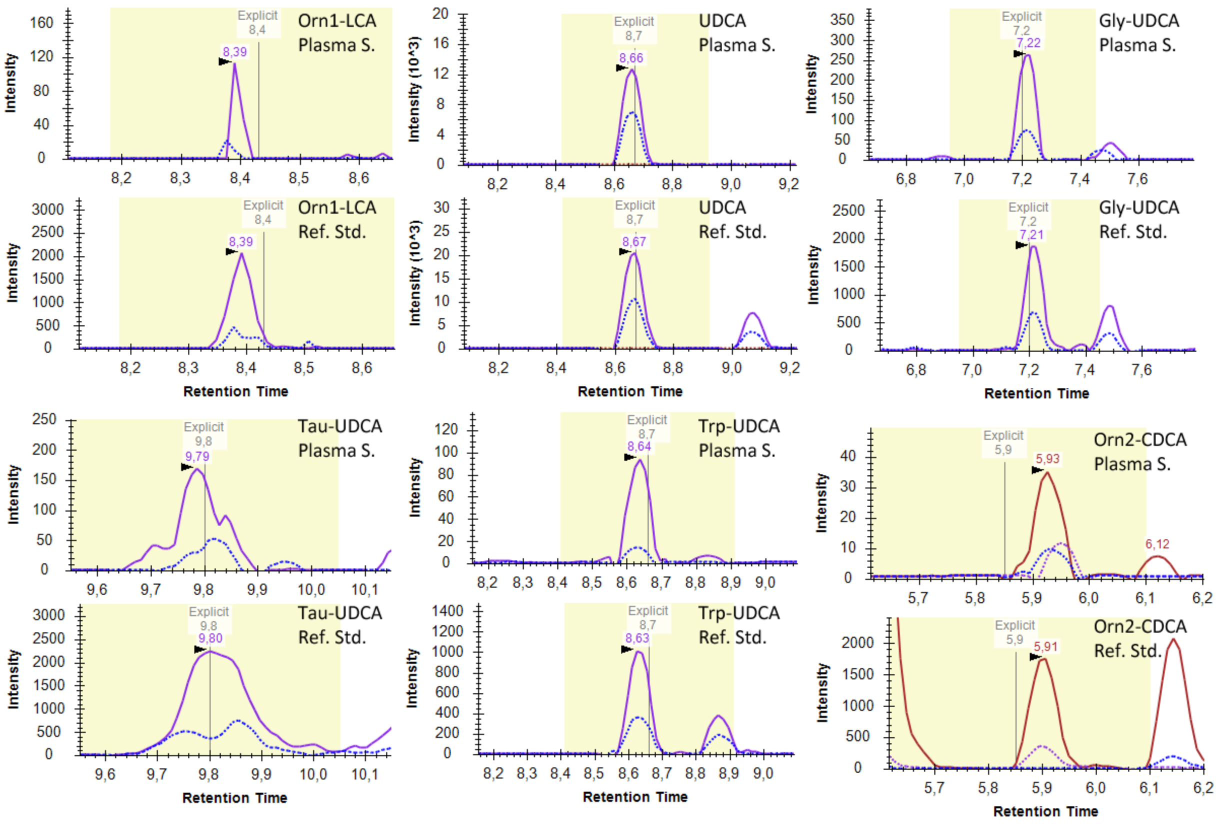

**Fig. S4.** (above) Extracted ion chromatograms of bile acids and bile acid conjugates detected in plasma samples (Plasma S.) compared to extracted ion chromatograms of the same compounds in the sample containing the reference standards (ref.Std.).

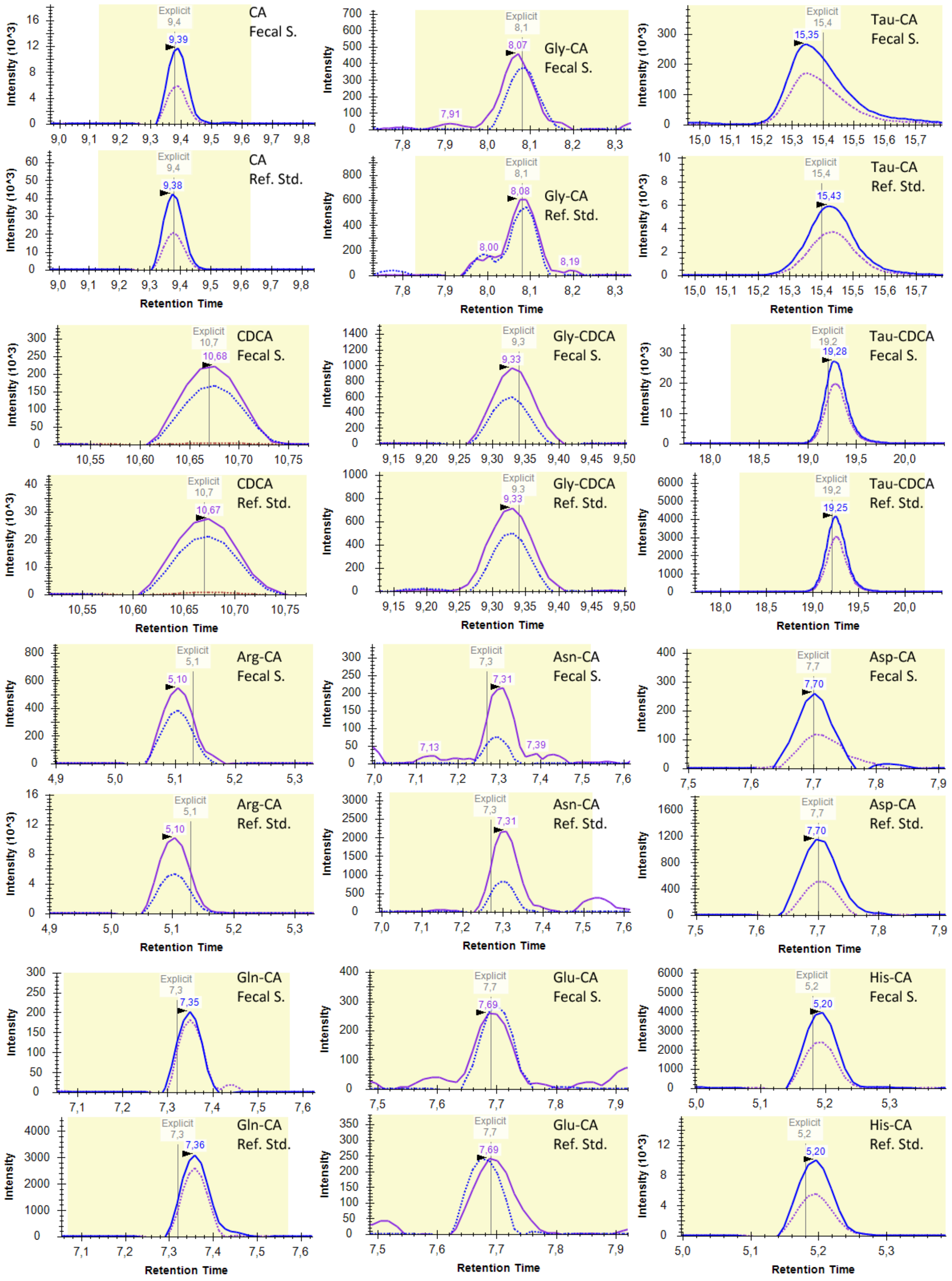
CA
Fecal S.
CA
Ref. Std.
Gly-CA
Fecal S.
Gly-CA
Ref. Std.
Tau-CA
Fecal S.
Tau-CA
Ref. Std.
CDCA
Fecal S.
CDCA
Ref. Std.
Gly-CDCA
Fecal S.
Gly-CDCA
Ref. Std.
Tau-CDCA
Fecal S.
Tau-CDCA
Ref. Std.
Arg-CA
Fecal S.
Arg-CA
Ref. Std.
Asn-CA
Fecal S.
Asn-CA
Ref. Std.
Asp-CA
Fecal S.
Asp-CA
Ref. Std.
Gln-CA
Fecal S.
Gln-CA
Ref. Std.
Glu-CA
Fecal S.
Glu-CA
Ref. Std.
His-CA
Fecal S.
His-CA
Ref. Std.
Intensity (10^3)
Intensity
Retention Time
Explicit

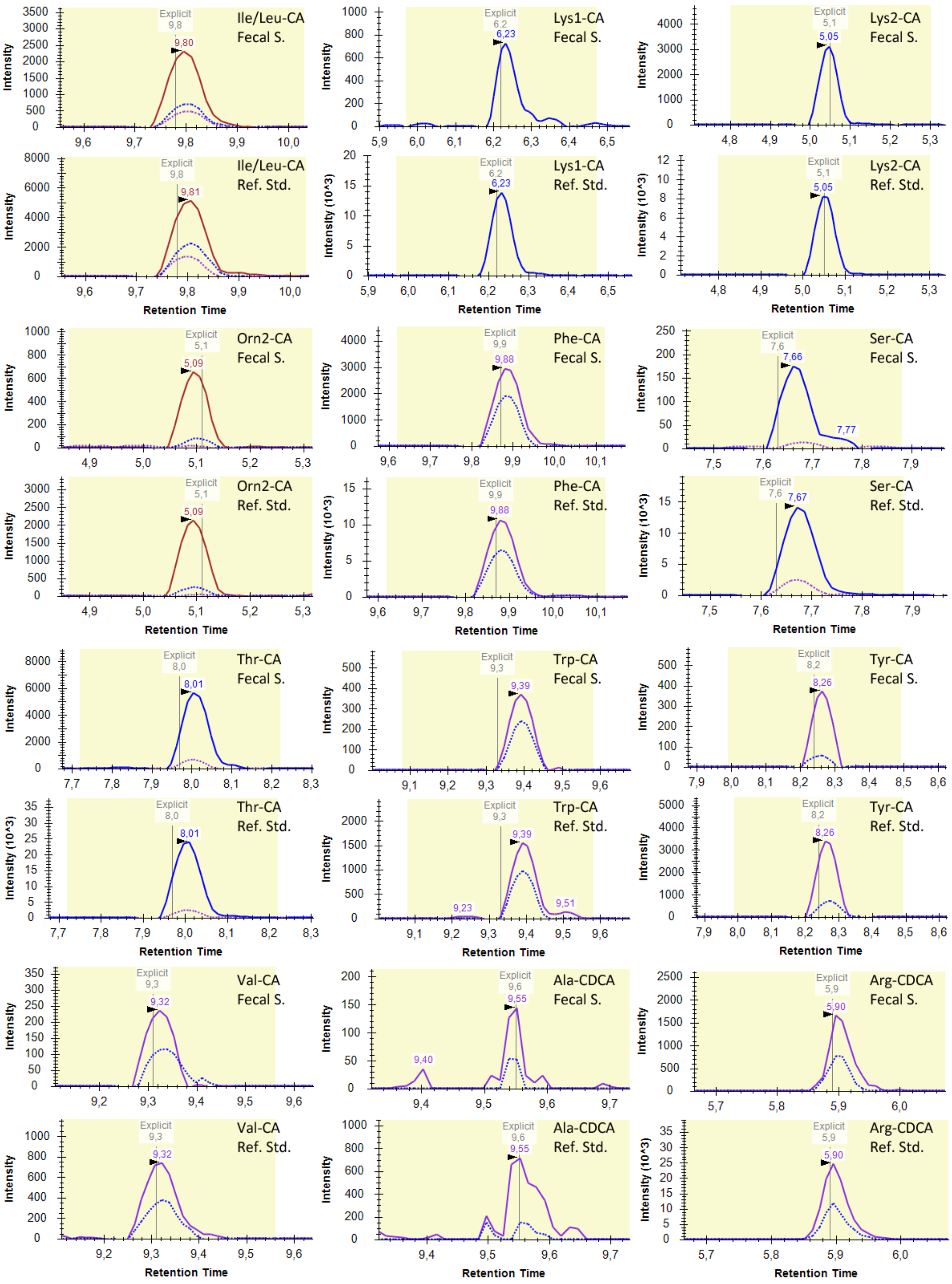

Ile/Leu-CA Fecal S.
Lys1-CA Fecal S.
Lys2-CA Fecal S.
Ile/Leu-CA Ref. Std.
Lys1-CA Ref. Std.
Lys2-CA Ref. Std.
Orn2-CA Fecal S.
Phe-CA Fecal S.
Ser-CA Fecal S.
Orn2-CA Ref. Std.
Phe-CA Ref. Std.
Ser-CA Ref. Std.
Thr-CA Fecal S.
Trp-CA Fecal S.
Tyr-CA Fecal S.
Thr-CA Ref. Std.
Trp-CA Ref. Std.
Tyr-CA Ref. Std.
Val-CA Fecal S.
Ala-CDCA Fecal S.
Arg-CDCA Fecal S.
Val-CA Ref. Std.
Ala-CDCA Ref. Std.
Arg-CDCA Ref. Std.
Intensity
Intensity (10^3)
Retention Time
Explicit

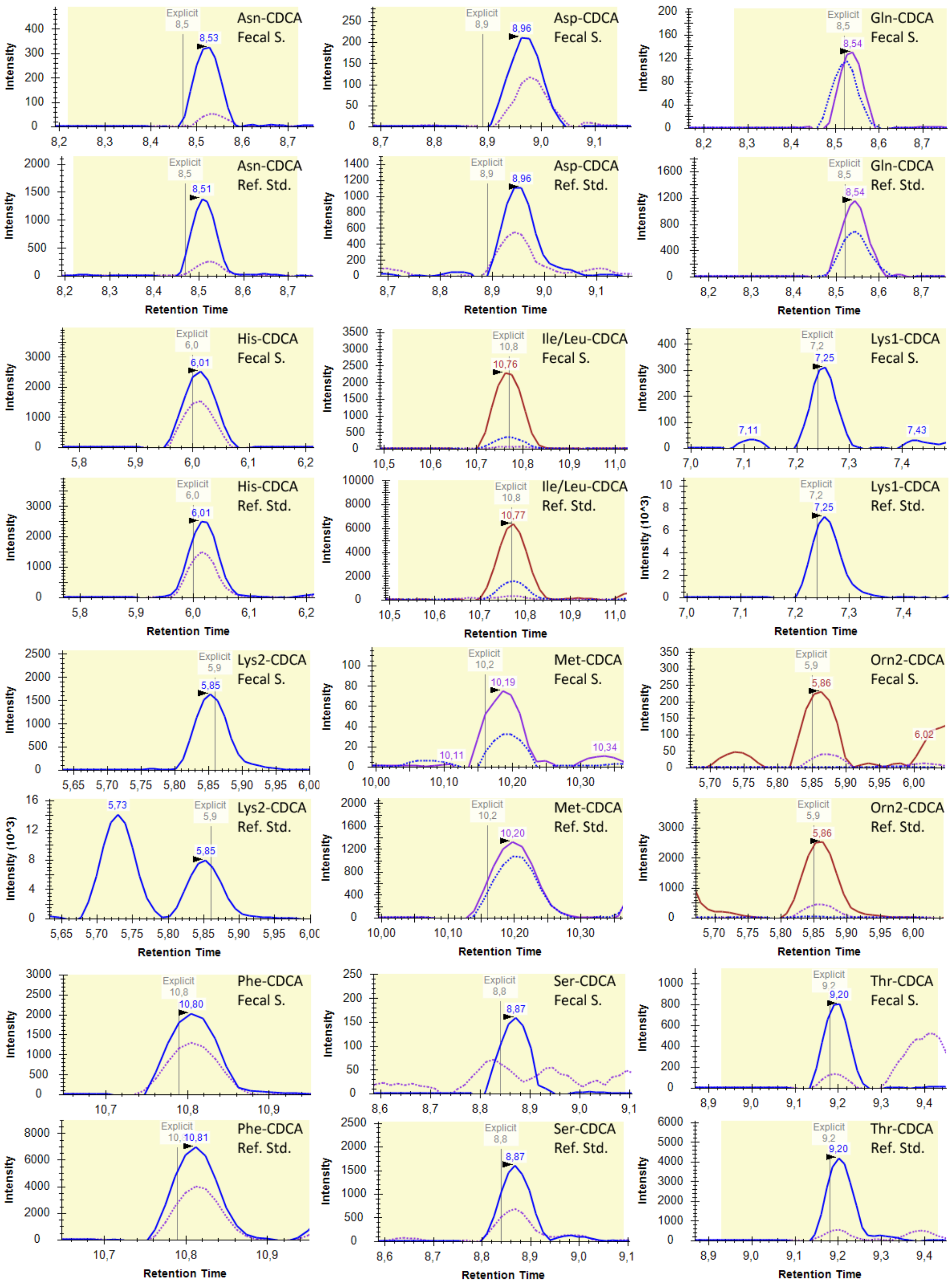
Asn-CDCA Fecal S.
Asp-CDCA Fecal S.
Gln-CDCA Fecal S.
Asn-CDCA Ref. Std.
Asp-CDCA Ref. Std.
Gln-CDCA Ref. Std.
His-CDCA Fecal S.
Ile/Leu-CDCA Fecal S.
Lys1-CDCA Fecal S.
His-CDCA Ref. Std.
Ile/Leu-CDCA Ref. Std.
Lys1-CDCA Ref. Std.
Lys2-CDCA Fecal S.
Met-CDCA Fecal S.
Orn2-CDCA Fecal S.
Lys2-CDCA Ref. Std.
Met-CDCA Ref. Std.
Orn2-CDCA Ref. Std.
Phe-CDCA Fecal S.
Ser-CDCA Fecal S.
Thr-CDCA Fecal S.
Phe-CDCA Ref. Std.
Ser-CDCA Ref. Std.
Thr-CDCA Ref. Std.
Intensity
Intensity (10^3)
Retention Time
Explicit

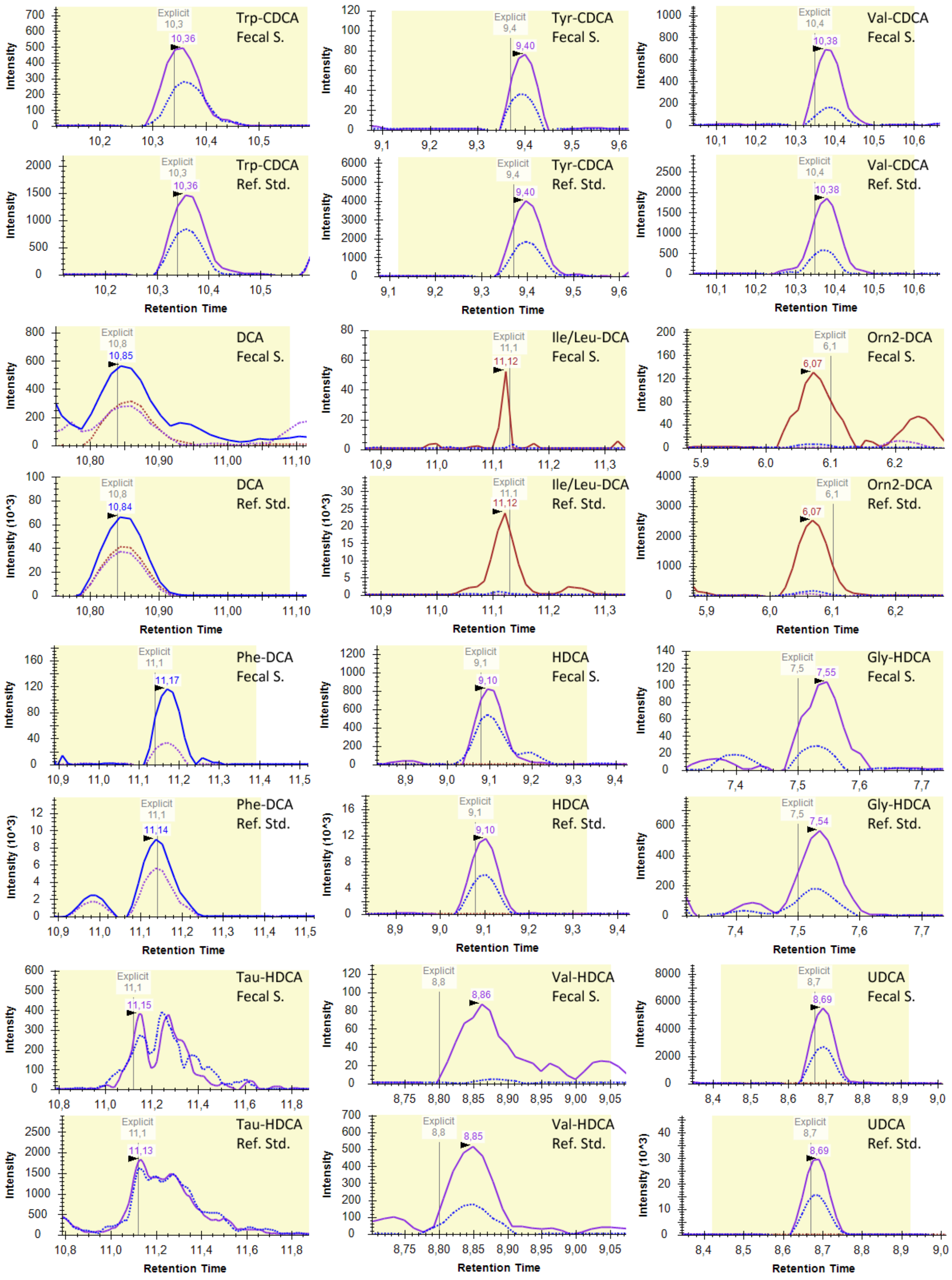
Trp-CDCA Fecal S.
Tyr-CDCA Fecal S.
Val-CDCA Fecal S.
Trp-CDCA Ref. Std.
Tyr-CDCA Ref. Std.
Val-CDCA Ref. Std.
DCA Fecal S.
Ile/Leu-DCA Fecal S.
Orn2-DCA Fecal S.
DCA Ref. Std.
Ile/Leu-DCA Ref. Std.
Orn2-DCA Ref. Std.
Phe-DCA Fecal S.
HDCA Fecal S.
Gly-HDCA Fecal S.
Phe-DCA Ref. Std.
HDCA Ref. Std.
Gly-HDCA Ref. Std.
Tau-HDCA Fecal S.
Val-HDCA Fecal S.
UDCA Fecal S.
Tau-HDCA Ref. Std.
Val-HDCA Ref. Std.
UDCA Ref. Std.
Intensity
Intensity (10^3)
Retention Time
Explicit

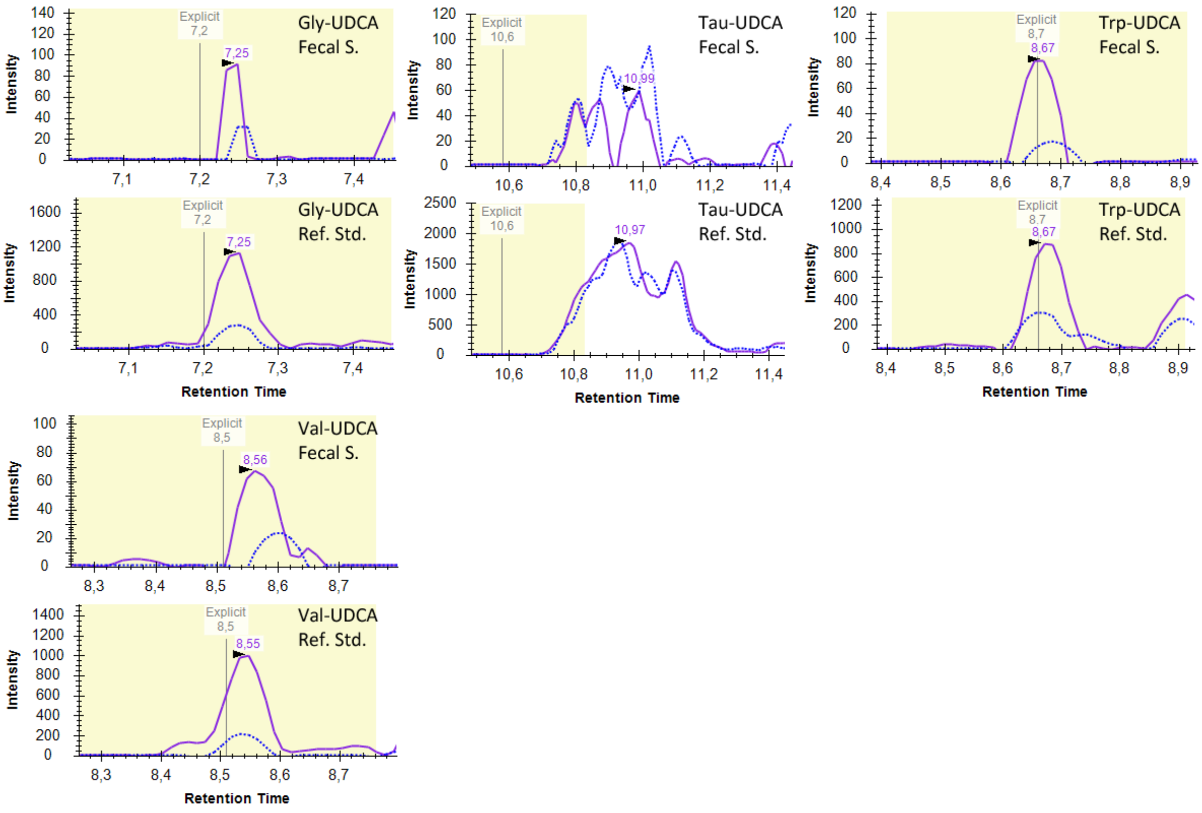

**Fig. S5.** (Above) Extracted ion chromatograms of bile acids and bile acid conjugates detected in fecal samples (Fecal S.) compared to extracted ion chromatograms of the same compounds in the sample containing the reference standards (Ref.Std.).

### 2 Additional plots and tables

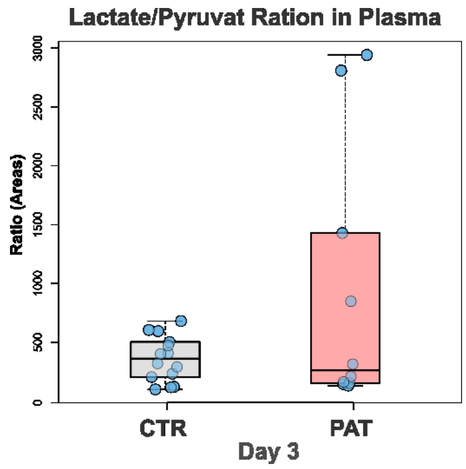

**Fig. S6.** Ratio of lactate and pyruvat in plasma at sampling time point day 3 in the control and pathological group.

**Tab. S3.** Selected compounds of interest in plasma with their $log_2$ fold change FC (PAT/CTR), p-value, identification confidence level (ICL)[33] and compound CAS number. A positive FC ($log_2$) indicates an increased abundance in infants exhibiting brain impairments (pathological group).

| Time point | Compound | FC ($log_2$) | p-value | q-value | ICL | CAS |
|---|---|---|---|---|---|---|
| **Plasma** | | | | | | |
| Day 3 | Alanine | 0.6 | 0.001 | 0.052 | 1 | 56-41-7 |
| Day 3 | Glutamine | 0.5 | 0.002 | 0.057 | 1 | 56-85-9 |
| Day 3 | Asparagine | 0.9 | 0.005 | 0.061 | 1 | 70-47-3 |
| Day 3 | Galactitol | 1.3 | 0.006 | 0.061 | 1 | 608-66-2 |
| Day 3 | Serine | 0.7 | 0.007 | 0.061 | 1 | 56-45-1 |
| Day 3 | Glycine | 0.7 | 0.019 | 0.086 | 1 | 56-40-6 |
| Day 3 | Guanidineacetic acid | 0.8 | 0.022 | 0.118 | 1 | 352-97-6 |
| Day 3 | Proline | 0.3 | 0.036 | 0.118 | 1 | 147-85-3 |
| Day 3 | Homoserine | 0.6 | 0.042 | 0.118 | 1 | 672-15-1 |
| Day 3 | Citrulline | 0.8 | 0.042 | 0.118 | 1 | 372-75-8 |
| Day 3 | Glyoxylic acid | 0.7 | 0.048 | 0.154 | 1 | 298-12-4 |
| Day 3 | Sarcosine | 1.3 | 0.048 | 0.121 | 1 | 107-97-1 |
| Day 3 | Indoleacrylic acid | 1.1 | 0.044 | 0.435 | 3 | 29953-71-7 |
| Day 3 | 3,5-Tetrahydroaldosterone sulfate | -1 | 0.007 | 0.032 | 3 | - |
| Day 3 | Tauro-1-hydroxycholic Acid | -3.1 | 0.006 | 0.032 | 3 | - |
| Day 3 | Dopamine 3-O-sulfate | -1.3 | 0.002 | 0.019 | 3 | 51317-41-0 |
| Day 3 | p-Phenolsulfonic acid | -0.8 | 0.024 | 0.052 | 3 | 585-38-6 |
| Day 3 | Phenol sulfate | -0.7 | 0.048 | 0.072 | 3 | 98-67-9 |
| Day 3 | 2-hydroxybenzenesulfonic acid | -1 | 0.048 | 0.072 | 3 | 937-34-8 |
| Day 3 | Cortisol 21-sulfate | -1.1 | 0.022 | 0.051 | 3 | 1253-43-6 |
| Day 3 | Succinate | 0.5 | 0.048 | 0.154 | 1 | 110-15-6 |
| Day 3 | Glycocholic acid | -1.2 | 0.11 | 0.106 | 1 | 475-31-0 |
| Day 3 | Taurocholic acid | -1.6 | 0.036 | 0.064 | 1 | 81-24-3 |
| Day 3 | Galactonic acid | -1.1 | 0.007 | 0.032 | 3 | 13382-27-9 |
| Day 3 | Inositol | -1.1 | 0.015 | 0.075 | 3 | 6917-35-7 |
| Day 3 | Tyrosol | 1.4 | 0.041 | 0.066 | 3 | 501-94-0 |
| Day 3 | Tryptamine | 1.7 | 0.012 | 0.104 | 1 | 61-54-1 |
| Day 7 | Pyruvate | -2.2 | 0.039 | 0.241 | 1 | 127-17-3 |
| Day 7 | Tryptamine | -1.4 | 0.048 | 0.161 | 1 | 61-54-1 |
| Day 7 | Indolelactic acid | 1 | 0.044 | 0.161 | 2.a | 87-51-4 |

| Time point | Compound | FC ($log_2$) | p-value | q-value | ICL | CAS |
|---|---|---|---|---|---|---|
| **Plasma** | | | | | | |
| Day 7 | Lanthionine | -1.1 | 0.039 | 0.161 | 3 | 922-55-4 |
| Day 7 | 3-Hydroxybenzoic acid | 1 | 0.002 | 0.265 | 3 | 99-06-9 |
| Day 28 | Pyruvate | 1.6 | 0.025 | 0.35 | 1 | 127-17-3 |
| Day 28 | Glyoxylic acid | 0.8 | 0.031 | 0.35 | 1 | 298-12-4 |
| Day 28 | *p*-cresol sulfate | 1.1 | 0.067 | 0.231 | 1 | 3233-58-7 |
| Day 28 | Dopamine | 1.1 | 0.067 | 0.231 | 1 | 51-61-6 |
| Day 28 | Indolpropionic acid | 0.8 | 0.11 | 0.278 | 1 | 830-96-6 |
| Day 28 | N-acetylcitrulline | -1.6 | 0.024 | 0.162 | 3 | 33965-42-3 |
| Day 28 | 3-Hydroxyoctanoic acid | 0.8 | 0.045 | 0.386 | 3 | 14292-27-4 |
| Day 28 | Succinylacetoacetate | 0.51 | 0.157 | 0.414 | 3 | 65115-74-4 |
| Week 32 | Dopamine 3-O-sulfate | -1.8 | 0.014 | 0.106 | 3 | 51317-41-0 |
| Week 32 | Cortisol 21-sulfate | -0.6 | 0.021 | 0.122 | 3 | 1253-43-6 |
| Term | *p*-cresol sulfate | -4.1 | 0.172 | 0.254 | 1 | 3233-58-7 |
| Term | 5-Oxo-delta-bilirubin | -1.4 | 0.013 | 0.091 | 3 | - |
| Term | Bilirubin IXα | -1.6 | 0.052 | 0.113 | 2.b | - |
| Term | Bilirubin IXβ | -1.5 | 0.038 | 0.104 | 1 | 635-65-4 |
| Term | Biliverdin IXα | -1 | 0.06 | 0.118 | 2.b | - |
| Term | Biliverdin IXβ | -1.4 | 0.027 | 0.096 | 1 | 114-25-0 |
| Term | Cortisol 21-sulfate | -1.1 | 0.007 | 0.034 | 3 | 1253-43-6 |
| Term | Estriol 3-sulfate | -1.5 | 0.023 | 0.049 | 3 | 481-95-8 |
| Term | 1,11-Undecanedicarboxylic acid | -1.2 | 0.005 | 0.032 | 3 | 505-52-2 |
| Term | Dodecanedioic acid | -1.3 | 0.013 | 0.032 | 3 | 693-23-2 |
| Term | 19-nortestosterone sulfate | -0.7 | 0.038 | 0.05 | 3 | - |
| Term | Tetrahydro-aldosteron sulfate | -0.4 | 0.023 | 0.067 | 3 | - |
| Term | 3-β,16α-Dihydroxyandrostenone sulfate | -1.1 | 0.045 | 0.109 | 3 | 4873-65-8 |
| Term | 4-Pyridoxic acid | -1.4 | 0.008 | 0.091 | 3 | 82-82-6 |
| Term | Undecanedioic acid | -1.1 | 0.001 | 0.034 | 3 | 1852-04-6 |
| Term | α-Aminoadipic acid | -1 | 0.027 | 0.147 | 1 | 1118-90-7 |
| Term | Indole | 1.2 | 0.022 | 0.094 | 1 | 120-72-9 |

**Tab. S4.** Selected compounds of interest in feces with their $log_2$ fold change FC (PAT/CTR), p-value, identification confidence level (ICL)[33] and compound CAS number. A positive FC ($log_2$) indicates an increased abundance in infants exhibiting brain impairments (pathological group).

| Time point | Compound | FC ($log_2$) | p-value | q-value | ICL | CAS |
|---|---|---|---|---|---|---|
| **Feces** | | | | | | |
| Day 7 | Tryptamine | 3.3 | 0.269 | 0.317 | 1 | 61-54-1 |
| Day 7 | Methionine sulfone | -4.5 | 0.013 | 0.174 | 1 | 7314-32-1 |
| Day 7 | Uracil | 3.4 | 0.041 | 0.174 | 1 | 66-22-8 |
| Day 7 | Histamine | 7 | 0.368 | 0.352 | 3 | 51-45-6 |
| Day 7 | Indolelactic acid | 4.9 | 0.048 | 0.195 | 2.a | 87-51-4 |
| Day 7 | Epipregnenolone sulfate | 2.7 | 0.004 | 0.111 | 3 | - |
| Day 7 | Pregenolone Sulfate | 1.8 | 0.109 | 0.14 | 3 | 1247-64-9 |
| Day 7 | Corticosterone sulfate | 1.4 | 0.048 | 0.111 | 3 | 1105-02-8 |
| Day 7 | 3β,16α-Dihydroxyandrostenone sulfate | 1 | 0.073 | 0.117 | 3 | 4873-65-8 |
| Day 7 | 16α-Hydroxy DHEA 3-sulfate | 1.2 | 0.073 | 0.117 | 3 | 4873-65-8 |
| Day 7 | Isonicotinic acid | 2.3 | 0.016 | 0.208 | 2.a | 55-22-1 |
| Day 7 | Adipic acid | 4.7 | 0.073 | 0.195 | 3 | 124-04-9 |
| Day 7 | α-Phocaecholic acid | 4 | 0.004 | 0.111 | 3 | 6879-45-4 |
| Day 7 | Indoleacetic acid | 2.4 | 0.153 | 0.236 | 1 | 87-51-4 |
| Day 7 | Chenodeoxycholic acid | 3.3 | 0.033 | 0.111 | 1 | 474-25-9 |
| Day 7 | Taurochenodeoxycholic acid | -3 | 0.067 | 0.116 | 1 | 516-35-8 |
| Day 7 | Cholic acid | 1.6 | 0.11 | 0.14 | 1 | 81-25-4 |
| Day 7 | Taurocholic acid | -3 | 0.11 | 0.14 | 1 | 81-24-3 |
| Day 28 | Dopamine | -0.8 | 0.042 | 0.183 | 1 | 51-61-6 |
| Day 28 | Indol-3-carbaldehyd | 0.8 | 0.092 | 0.254 | 1 | 487-89-8 |
| Day 28 | Tyrosine | 2.2 | 0.078 | 0.366 | 1 | 60-18-4 |
| Day 28 | 5-Hydroxyindole | -2.7 | 0.005 | 0.183 | 3 | 1953-54-4 |
| Day 28 | Adipic acid | 1.7 | 0.004 | 0.154 | 3 | 124-04-9 |
| Day 28 | N-Acetylneuraminic acid | 0.6 | 0.191 | 0.29 | 3 | 131-48-6 |
| Term | Dopamine | 1.3 | 0.018 | 0.583 | 1 | 51-61-6 |
| Term | Proline | 0.9 | 0.03 | 0.201 | 1 | 147-85-3 |
| Term | Alanine | 0.8 | 0.03 | 0.201 | 1 | 56-41-7 |

### 3 Compound identification

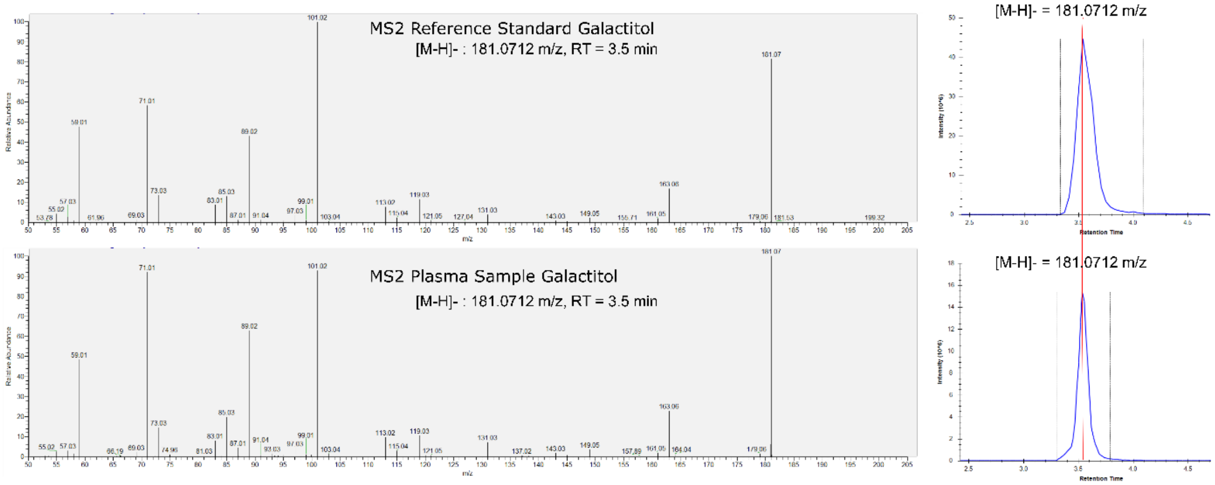

**Fig. S7.** $MS^2$ fragmentation pattern and extracted ion chromatogram of galactitol reference standard (2 µM) compared to feature found in biological sample and identified as galactitol.
